# Loss of a Spouse and Risk of Cognitive Decline: Insights from Six Prospective Cohort Studies

**DOI:** 10.64898/2026.05.20.26353668

**Authors:** C. Guo, Y. Wang, X. Sun, F. Ge

## Abstract

**Aims:** The risk of cognitive decline after losing a spouse remained mixed. This study aims to investigate the association between spousal loss and risk of cognitive decline, assess whether this association varies by sex and age, and identify modifiable factors.

**Methods:** We conducted a prospective cohort study using harmonized data from six population-based aging surveys: the US Health and Retirement Study and its sister surveys in England, Mexico, China, India, and South Africa, incorporating their respective Harmonized Cognitive Assessment Protocol (HCAP) sub-studies. Spousal loss (yes vs no) was the exposure. Cognitive outcomes (i.e., orientation, memory, executive function, and language), were assessed using HCAP neuropsychological batteries. We conducted parallel analyses in six cohorts. Associations between spousal loss and cognitive outcomes were estimated using generalized linear models, and summarised estimates were derived via random-effects meta-analyses. Sex stratification and restricted cubic spines were used to examine how these associations vary by sex and age, respectively.

**Results:** The analytical cohort consisted of 18,551 individuals aged 61.22 (SD 6.30) to 71.37 (SD 7.33) years. Widowhood prevalence ranged from 14.1% in CHARLS to 53.9% in HAALSI and was consistently higher in women. Spousal loss was associated with poorer memory (multivariable-adjusted *β* = -0.07, 95% CI -0.12 to -0.01) and executive function (multivariable-adjusted *β* = -0.08, 95% CI -0.13 to -0.03) in the meta-analysis, with no significant associations for orientation or language. While results were generally consistent in five cohorts, the ELSA showed divergent patterns (orientation: *β* = 0.10, 95% CI 0.06 to 0.13; memory: *β* = 0.05, 95% CI 0.02 to 0.08; language: *β* = 0.16, 95% CI 0.12 to 0.19). Sex-stratified analyses indicated poorer executive function among men (multivariable-adjusted *β* = -0.14, 95% CI -0.19 to -0.08) and poorer memory among women (multivariable-adjusted *β* = -0.07, 95% CI -0.14 to -0.01) following widowhood. Nonlinear age-related effects on cognition were observed in ELSA, LASI, and HAALSI. Higher education, internet use, and BMI were negatively associated with the risk of cognitive decline among widowed participants.

**Conclusions:** Spousal loss is associated with domain- and sex-specific differences in cognitive performance, with substantial heterogeneity across study populations. Future research should integrate biopsychosocial markers to develop context-sensitive interventions for widowed older adults.

## 1. Introduction

As the global population ages, the incidence of age-related diseases, particularly dementia, is on the rise, placing increasing pressure on individuals, families, and healthcare systems (Patterson 2018, Livingston et al. 2024). Currently, approximately 57 million people are living with dementia, with about two-thirds residing in low- and middle-income countries, and this number is projected to exceed 153 million by 2050 (GBD 2019 Dementia Forecasting Collaborators 2022). Cognitive decline, an early hallmark of dementia, plays a crucial role in facilitating early detection and enabling timely intervention. In response to the growing burden of aging, large-scale aging studies such as the US Health and Retirement Study (HRS) and its international sister studies have been established, alongside the Harmonized Cognitive Assessment Protocol (HCAP) sub-studies, which standardizes cognitive measures across countries to enable cross-national comparisons (Langa et al. 2020, Gross et al. 2023).

Previous studies have shown that exposure to traumatic life events is associated with an increased risk of dementia (Severs et al. 2023), and emerging evidence suggests that the loss of a spouse constitutes a negative late-life stressor with potential cognitive consequences. The stress model (Williams & Umberson 2004) posits that spousal loss constitutes a life stressor requiring substantial adaptation, with sustained stress adversely affecting mental health. Meanwhile, the marital resource model (Waite et al. 2001) emphasizes that marriage provides social, psychological, and economic resources that protect physical and mental health. Accordingly, widowed older adults—particularly those with limited socioeconomic resources—are expected to exhibit poorer cognitive function.

Notwithstanding the growing recognition of associations between widowhood and subsequent cognitive changes, much of the existing evidence comes from high-income countries and remains inconsistent. For example, some studies have reported increased risks of cognitive decline and dementia (Håkansson et al. 2009, Sommerlad et al. 2018, Shin et al. 2018, Zhang et al. 2019, Liu et al. 2019, 2020, Singham et al. 2021, Min & Song 2023), whereas others have found no significant association (Vidarsdottir et al. 2014, Lee et al. 2019, Xu et al. 2021, Nerobkova et al. 2022, Lee & Jiang 2023). Thus, a comprehensive study that includes both developed and developing countries is essential for improving our understanding of these associations.

Indeed, previous studies on widowhood have largely focused on psychosocial factors like pre-loss marital quality (Chen et al. 2022), whereas cognitive decline is a multifactorial condition influenced by a variety of risk factors, such as educational attainment and health-related behaviors (Zhang et al. 2024, Livingston et al. 2024). Therefore, additional research is needed to investigate other potential factors that underlie the association between widowhood and cognitive health.

Using data from six countries, this study aims (1) to examine whether spousal loss is differentially associated with specific cognitive domains, including orientation, memory, executive function, and language; (2) to assess whether these associations vary by sex and age; and (3) to identify individual-level characteristics associated with better cognitive performance among widowed individuals.

## 2. Methods

### 2.1 Data sources and study design

This observational prospective study included six cohorts from six countries (United States, England, Mexico, China, India, and South Africa). These cohorts are all part of the Gateway to Global Aging Data project (https://g2aging.org/), described elsewhere (Lee et al. 2021). The cohorts were the HRS, English Longitudinal Study of Ageing (ELSA), the Mexican Health and Aging Study (MHAS), the China Health and Retirement Longitudinal Study (CHARLS), the Longitudinal Ageing Study in India (LASI), and the Health and Aging in Africa: A Longitudinal Study of an INDEPTH Community in South Africa (HAALSI). All cohorts had ethical approvals from respective national or regional ethics committees with varying numbers of waves of data collections. All participants provided written or electronic informed consent. Details of these cohorts are shown in the Fig. 1A and Supplementary information.

### 2.2 Measures

#### 2.2.1 Procedures

The six cohorts had varying numbers of waves, ranging from two to four. To minimize reverse causality between the exposure (spousal loss) and outcomes (cognitive function), we used the earlier waves to define spousal loss and then used cognitive function data from the subsequent wave. For three cohorts (HRS, ELSA, and CHARLS), spousal loss was defined using waves 1 to 3, and cognitive function data from wave 4 were used (Fig. 1B and 1C). Detailed information on the waves used to define spousal loss in each cohort is shown in Fig. S1–S6.

#### 2.2.2 Ascertainment of Spousal loss (Exposure)

We defined the exposure and control based on the previous studies (Lee et al. 2019). Spousal loss was defined using self-reported legal marital status. Detailed information on how marital status was measured in six cohorts is shown in Table S1. Prior evidence indicates that legal marriage confers greater social and legal stability than cohabitation and is associated with more favorable emotional and cognitive outcomes (Perelli Harris & Styrc 2018), therefore, participants who were partnered but not legally married (i.e., cohabiting) were excluded from the primary analyses. The exposure group included individuals who were widowed at baseline as well as those who reported widowhood at subsequent waves, provided they did not remarry or repartner thereafter. The control group consisted of participants who remained continuously married throughout the observation period.

#### 2.2.3 Ascertainment of cognitive function (Outcome)

Given that previous mixed findings may be due to the different cognitive outcomes examined (e.g., global cognitive function, Alzheimer’s disease, and dementia), this study assessed cognitive function using the HCAP neuropsychological battery from six countries. We used standardized *z* scores of domain-specific cognitive functions (orientation, memory, executive function, and language) as the primary outcome in this study. Detailed information on the measurement of cognitive function is shown in Table S2.

#### 2.2.4 Covariates

Covariates were selected based on established theoretical frameworks, with primary reference to the 14 modifiable dementia risk factors identified by the 2024 Lancet Commission (Livingston et al. 2024), while also considering data availability and cross-cohort harmonization. All selected covariates included age (continuous in year), sex (male or female), minority status [ethnicity (white/caucasian or non-white) or living region (urban community or rural village)], educational attainment (less than upper secondary, upper secondary/vocational training or tertiary), household wealth level (low, normal or high), BMI (underweight/normal, overweight or obesity), smoking status (current, former or never), drinking patterns (heavy, moderate, light or never), internet use (no or yes), comorbidities (mild, moderate or severe), and baseline cognition (≤25%, 25∼50%, 50%∼75% or >75%). Detailed information on the measurement of these covariates is provided in Table S3.

#### 2.2.5 Statistical analysis

First, we examined the distribution of sociodemographic and health-related factors between widowed and married groups within each cohort, using means with standard deviations (SD) for continuous variables and counts with percentages for categorical variables.

Associations between spousal loss and domain-specific cognitive function were examined using generalized linear models, with results reported as *β* coefficients and 95% confidence intervals (95% CI). To address whether these associations varied by sex, analyses were conducted separately for males and females. All models were adjusted for a common set of covariates defined a priori. We meta-analysed based on effect size that estimated from each cohort with a random-effects model using the metafor package in R to estimate overall *β* coefficients (Viechtbauer 2010). Heterogeneity for each overall cognitive outcomes was examined using the *I*² statistic (Higgins et al. 2003).

To account for potential nonlinear age-related cognitive changes associated with widowhood (Shin et al. 2018, Liu et al. 2019), we applied restricted cubic spline models with four knots and incorporated an age × widowhood interaction to assess effect modification by widowhood status.

Finally, we conducted analyses restricted to widowed individuals to estimate the associations between the aforementioned covariates and cognitive outcomes. Multivariable generalized linear models were used to estimate *β* coefficients, adjusting for age, sex, minority status, and baseline cognition.

We performed several sensitivity analyses. First, to assess the effect of missingness on our results, we re-ran our analysis restricted to participants without the missing values. Second, to address potential concerns regarding temporal ordering and reverse causation between widowhood and cognitive function, we conducted an analysis restricting the sample to baseline participants only. Third, considering the strong influence of aging on cognitive decline, we further restricted the HCAP sample to participants aged 65 years and older and re-evaluated the analysis. Last, to examine whether the meta-analytic results were disproportionately driven by high-income countries, we conducted an additional sensitivity analysis excluding results estimated from HRS and ELSA and re-ran the meta-analyses using the remaining four cohorts.

All analyses were conducted in Stata/MP 18.0 and R 4.3.2, with statistical significance defined as a two-sided *P* value < 0.05. Further details are provided in the Supplementary Methods.

## 3. Results

### 3.1 Participant characteristics

A total of 18,551 participants were included from six cohorts (Table 1). The baseline mean age (SD) ranged from 61.22 (6.30) to 71.37 (7.33) years. Specifically, 2,672 participants from HRS, 1,075 from ELSA, 1,553 from MHAS, 8,757 from CHARLS, 3,924 from LASI and 570 from HAALSI. The proportion of widowed individuals varied substantially, ranging from 14.1% in CHARLS to 53.9% in HAALSI, with a consistently higher prevalence among female participants. Detailed descriptive information for widowed and married individuals in each cohort is shown in Tables S4-S9.

**Table 1:** Characteristics of participants by countries.

|  | **HRS**<br>**N=2672** | **ELSA**<br>**N=1075** | **MHAS**<br>**N=1553** | **CHARLS**<br>**N=8757** | **LASI**<br>**N=3924** | **HAALSI**<br>**N=570** |
| --- | --- | --- | --- | --- | --- | --- |
| **Widowed group** | 851 (31.9) | 308 (28.7) | 407 (26.2) | 1231 (14.1) | 1313 (33.5) | 307 (53.9) |
| **Age, mean (SD), y** | 70.57 (7.50) | 71.06 (7.22) | 65.35 (8.38) | 61.39 (6.40) | 68.86 (7.43) | 64.99 (11.29) |
| **Minority status** | 475 (17.8) | 31 (2.9) | 469 (30.2) | 5254 (60.0) | 2453 (62.5) | NA |
| **Sex** |  |  |  |  |  |  |
| Male | 1077 (40.3) | 475 (44.2) | 656 (42.2) | 4446 (50.8) | 1794 (45.7) | 201 (35.3) |
| Female | 1595 (59.7) | 600 (55.8) | 897 (57.8) | 4311 (49.2) | 2130 (54.3) | 369 (64.7) |
| **Educational attainment** |  |  |  |  |  |  |
| Less than upper secondary | 461 (17.3) | 380 (35.3) | 1367 (88.0) | 7839 (89.5) | 2967 (75.6) | 524 (91.9) |
| Upper secondary or vocational training | 1543 (57.7) | 469 (43.6) | 39 (2.5) | 782 (8.9) | 800 (20.4) | 37 (6.5) |
| Tertiary | 667 (25.0) | 126 (11.7) | 132 (8.5) | 136 (1.6) | 157 (4.0) | 9 (1.6) |
| Missing | 1 (0.0) | 100 (9.3) | 15 (1.0) | 0 | 0 | 0 |
| **Household wealth level** |  |  |  |  |  |  |
| Low | 896 (33.5) | 357 (33.2) | 614 (39.5) | 3166 (36.2) | 1231 (31.4) | 169 (29.6) |
| Normal | 892 (33.4) | 357 (33.2) | 569 (36.6) | 2567 (29.3) | 1233 (31.4) | 167 (29.3) |
| High | 884 (33.1) | 357 (33.2) | 370 (23.8) | 2845 (32.5) | 1229 (31.3) | 167 (29.3) |
| Missing | 0 | 4 (0.4) | 0 | 179 (2.0) | 231 (5.9) | 67 (11.8) |
| **BMI** |  |  |  |  |  |  |
| Underweight/Normal | 495 (18.5) | 244 (22.7) | 469 (30.2) | 5605 (64.0) | 2563 (65.3) | 206 (36.1) |
| Overweight | 871 (32.6) | 444 (41.3) | 639 (41.1) | 2328 (26.6) | 783 (20.0) | 176 (30.9) |
| Obesity | 1090 (40.8) | 342 (31.8) | 389 (25.0) | 362 (4.1) | 255 (6.5) | 154 (27.0) |
| Missing | 216 (8.1) | 45 (4.2) | 56 (3.6) | 462 (5.3) | 323 (8.2) | 34 (6.0) |
| **Smoking status** |  |  |  |  |  |  |
| Current | 221 (8.3) | 95 (8.8) | 169 (10.9) | 2796 (31.9) | 1317 (33.6) | 32 (5.6) |
| Former | 1191 (44.6) | 590 (54.9) | 414 (26.7) | 870 (9.9) | 259 (6.6) | 61 (10.7) |
| Never | 1242 (46.5) | 390 (36.3) | 970 (62.5) | 5066 (57.9) | 2325 (59.3) | 477 (83.7) |
| Missing | 18 (0.7) | 0 | 0 | 25 (0.3) | 23 (0.6) | 0 |
| **Drinking patterns** |  |  |  |  |  |  |
| Heavy | 299 (11.2) | 239 (22.2) | 27 (1.7) | 1402 (16.0) | 54 (1.4) | 27 (4.7) |
| Moderate | 683 (25.6) | 408 (38.0) | 209 (13.5) | 400 (4.6) | 58 (1.5) | 30 (5.3) |
| Light | 468 (17.5) | 238 (22.1) | 134 (8.6) | 1802 (20.6) | 467 (11.9) | 201 (35.3) |
| Never | 1222 (45.7) | 161 (15.0) | 1183 (76.2) | 5130 (58.6) | 3324 (84.7) | 312 (54.7) |
| Missing | 0 | 29 (2.7) | 0 | 23 (0.3) | 21 (0.5) | 0 |
| **Internet use** |  |  |  |  |  |  |
| No | 1288 (48.2) | 479 (44.6) | 660 (42.5) | 7024 (80.2) | 3776 (96.2) | 74 (13.0) |
| Yes | 1384 (51.8) | 567 (52.7) | 893 (57.5) | 193 (2.2) | 96 (2.4) | 457 (80.2) |
| Missing | 0 | 29 (2.7) | 0 | 1540 (17.6) | 52 (1.3) | 39 (6.8) |
| **Comorbidities** |  |  |  |  |  |  |
| Mild | 1791 (67.0) | 816 (75.9) | 1172 (75.5) | 7395 (84.4) | 3313 (84.4) | 472 (82.8) |
| Moderate | 639 (23.9) | 212 (19.7) | 309 (19.9) | 751 (8.6) | 536 (13.7) | 57 (10.0) |
| Severe | 242 (9.1) | 47 (4.4) | 72 (4.6) | 151 (1.7) | 66 (1.7) | 7 (1.2) |
| Missing | 0 | 0 | 0 | 460 (5.3) | 9 (0.2) | 34 (6.0) |
| **Baseline cognition** |  |  |  |  |  |  |
| ≤25% | 856 (32.0) | 339 (31.5) | 396 (25.5) | 2906 (33.2) | 1082 (27.6) | 218 (38.2) |
| 25~50% | 685 (25.6) | 253 (23.5) | 381 (24.5) | 2059 (23.5) | 1236 (31.5) | 75 (13.2) |
| 50%~75% | 595 (22.3) | 235 (21.9) | 411 (26.5) | 1853 (21.2) | 742 (18.9) | 180 (31.6) |
| > 75% | 536 (20.1) | 248 (23.1) | 365 (23.5) | 1831 (20.9) | 820 (20.9) | 92 (16.1) |
| Missing | 0 | 0 | 0 | 108 (1.2) | 44 (1.1) | 5 (0.9) |

Unless indicated otherwise, data are expressed as No. (%) of participants. Percentages have been rounded and may not total 100.
HRS, the Health and Retirement Study; ELSA, the English Longitudinal Study of Ageing; MHAS, the Mexican Health and Aging Study; CHARLS, the China Health Retirement Longitudinal Study; LASI, the Longitudinal Aging Study in India; HAALSI, the Health and Aging in Africa: A Longitudinal Study of an INDEPTH Community in South Africa; SD, standard deviations; NA, not applicable; BMI, body mass index.

### 3.2 Spousal loss and cognitive function

Fig. 2 presents the *β* coefficients for the associations between spousal loss and cognitive outcomes in each cohort and in the meta-analysis. The meta-analysis showed that, compared with married individuals, widowhood was associated with poorer memory (multivariable-adjusted *β* = -0.07, *I*² = 98.27%) and poorer executive function (multivariable-adjusted *β* = -0.08, *I*² = 98.37%), but not with orientation (multivariable-adjusted *β* = -0.03, *I*² = 98.05%) or language (multivariable-adjusted *β* = -0.03, *I*² = 99.30%) (Table S10). The association patterns were broadly consistent in five cohorts, although the magnitude of the effect sizes varied. In contrast, data from ELSA showed a divergent pattern, with spousal loss associated with neutral or even better cognitive performance in several domains (orientation: *β* = 0.10, 95% CI 0.06 to 0.13; memory: *β* = 0.05, 95% CI 0.02 to 0.08; language: *β* = 0.16, 95% CI 0.12 to 0.19).

Fig. 3 presents the *β* coefficients for the associations between spousal loss and cognitive outcomes stratified by sex. Among male participants, widowhood was associated with poorer executive function (multivariable-adjusted *β* = -0.14, *I*² = 94.31%), but not with orientation (multivariable-adjusted *β* = 0, *I*² = 95.21%), memory (multivariable-adjusted *β* = -0.03, *I*² = 93.97%), or language (multivariable-adjusted *β* = -0.08, *I*² = 98.41%) (Table S11). Among female participants, widowhood was associated with poorer memory (multivariable-adjusted *β* = -0.07, *I*² = 97.65%), but not with orientation (multivariable-adjusted *β* = -0.03, *I*² = 97.15%), executive function (multivariable-adjusted *β* = -0.05, *I*² = 98.12%), or language (multivariable-adjusted *β* = -0.01, *I*² = 98.38%) (Table S12). Similar to the combined analyses of male and female participants, the ELSA cohort continued to show a divergent pattern in the sex-stratified analyses.

Fig. S7 and S8 reveal several nonlinear associations between age and cognitive performance, which varied by cohort and widowhood status. Significant nonlinear age-related patterns were observed for orientation, executive function, and language in LASI, as well as for executive function in ELSA. In addition, a borderline nonlinear trend was noted for memory in HAALSI (*P* = 0.05). However, evidence of effect modification by widowhood status was limited; a statistically significant age × widowhood interaction was detected only for executive function in ELSA (*P* = 0.04).

When specifically studying widowed individuals, we found that that higher educational attainment and internet use were consistently associated with better cognitive outcomes in multiple cohorts (Fig. 4; Tables S13–S18). In middle-income national cohorts such as LASI and MHAS, higher BMI was associated with better performance in executive function and language, with similar trends observed in CHARLS. Certain individual characteristics showed cohort-specific associations with cognitive outcomes among widowed individuals, such as a positive association between household wealth level and cognitive function in HRS and an adverse association between smoking and cognitive performance in HAALSI.

### 3.3 Sensitivity analyses

Associations remained in the same direction, though few reached statistical significance in the complete-case analysis, likely due to the substantially reduced sample size (Table S19). The estimates from the sensitivity analyses restricted to baseline participants (Table S20) or those aged 65 years and older in HCAPs (Table S21) were largely consistent to those from the main analysis, especially among male participants. In sensitivity analyses excluding HRS and ELSA, associations between widowhood and cognitive performance became more consistent. The pooled association for orientation strengthened markedly with minimal heterogeneity (multivariable-adjusted *β* = -0.08, *I*² = 1.97%), while associations for memory and executive function remained directionally consistent with slightly larger effect sizes (Table S22). And the association with language became statistically significant after exclusion of the two high-income national cohorts.

## 4. Discussion

The present multinational study represents one of the most comprehensive investigations to date examining the association between spousal loss and cognitive changes in diverse regions and socioeconomic contexts. By analyzing harmonized data from six prospective cohorts spanning high-, middle-income countries, we found that widowhood was associated with poorer cognitive performance in specific domains, particularly memory and executive function, with clear sex-specific patterns. The substantial cross-national heterogeneity, driven in part by divergent findings in ELSA, highlights the context-dependent nature of the cognitive consequences of spousal loss. In addition, we observed that age-related patterns of cognitive decline differed by widowhood status in a few cohorts, and that certain individual characteristics such as higher education, internet use, and BMI may mitigate the cognitive risks associated with spousal loss. Taken together, these findings suggest that the cognitive consequences of spousal loss are not driven by a single mechanism but reflect the interplay of biological vulnerability, psychological adaptation to loss, and social and contextual resources. This integrative framework provides a conceptual basis for developing context-sensitive, biopsychosocial interventions to support cognitive resilience among vulnerable subgroups of widowed older adults, particularly in resource-limited settings.

Our findings add to existing mixed literature by showing that, although spousal loss is associated with cognitive decline in many settings, these effects are domain-specific, sex-dependent, and vary from countries. A consistent downward trend in executive function was observed in nearly all regions, with more pronounced among men. Executive function—including working memory, cognitive flexibility, and inhibitory control—is critical for attention regulation and decision-making (Guevara et al. 2019), and appears particularly vulnerable to external stressors (Stroebe & Stroebe 1983). Impairment in executive function can lead to difficulties in daily function, which may partly explain the higher rates of functional limitations among widowed individuals compared with their married counterparts (Johnson et al. 2007, Hughes & Waite 2009). Evidence from one preliminary study further shows that bereaved spouses exhibit declines in working memory and cognitive inhibition (Wu-Chung et al. 2025). The marital resource model (Waite et al. 2001) may partly explain the observed sex-specific differences, particularly the consistent executive decline among widowed men. Men may rely more on their spouses for emotional regulation and instrumental support in daily life, including health management and household responsibilities (Umberson 1992, Kalmijn 2007). In contrast, widowed women are more likely to experience memory impairment. Previous evidence indicates that widowed women experience higher levels of grief and depression following spousal death (Chen et al. 1999), which can induce fluctuations in estrogen levels (Barth et al. 2023). These hormonal changes may impair memory-related brain regions such as the hippocampus. However, Aartsen et al. (2005) found that only widowed men, but not women, had a greater memory decline than their married counterparts. And the rate of memory decline over time was similar for widowed men and women. Orientation and language abilities tend to be relatively preserved in normal aging and are less sensitive to psychosocial stressors such as spousal loss compared with other cognitive domains, reflecting domain-specific patterns in cognitive aging(Rut et al. 2018).

Consistent with previous studies (Feng et al. 2014, Liu et al. 2020), widowhood is associated with sex differences in cognitive decline. In traditional marital pattern, women typically serve as family kin-keepers, providing emotional support to their husbands; therefore, losing a spouse tends to have a greater negative impact on men’s social networks and support systems (Williams & Umberson 2004), which are closely linked to cognitive impairment. Also, men tend to exhibit higher pre-bereavement engagement in harmful behaviors like excessive drinking. U.S. CDC data indicate that 25.6% of older men (vs. 12.4% of women) consume ≥4 drinks daily, a known risk factor for accelerated cognitive deterioration (Robin & Michael 2022). In countries with limited formal support systems and enduring gender inequalities—such as China, India, and South Africa—spousal loss may trigger more disruptions in social identity, access to care, and economic security. For instance, widowed women in India are often subjected to restrictive norms, including wearing white and dietary limitations, that deepen economic dependency and social exclusion (Singh 2022). In China, spousal loss is significantly associated with loneliness among older adults (Yang & Gu 2021), supporting that Chinese widow showed severe language decline because of reduced linguistic stimulation. Countries with better welfare systems may offer buffering effects. In England, public healthcare and social support services may facilitate adaptation to bereavement through improved access to medical care, community programs, or volunteer opportunities (Alderwick et al. 2022). However, given the elevated mortality risk linked to widowhood (Wang et al. 2024), the widowed population from England may reflect ‘healthy survivors’ (Marshall J. & Donald R. 1996), potentially leading to an underestimation of widowhood’s adverse impact on cognitive decline. In Mexico, policies like non-contributory pensions have been shown to improve older adults’ well-being and may indirectly preserve cognitive health by alleviating financial strain (Aguila & Casanova 2020). Moreover, multigenerational living arrangements (Aguila et al. 2020), prevalent in Mexico, may offer additional protection against cognitive decline through increased social interaction. Future cross-national research is warranted to elucidate how cultural contexts and macro policies jointly shape the cognitive consequences of spousal loss.

Our findings do not fully support the hypothesis that widowhood-related cognitive patterns change nonlinearly with marital status (Shin et al. 2018, Liu et al. 2019). Although cognitive function generally declined with age, such declines followed a nonlinear trajectory among widowed individuals only in certain domains and cohorts. These limited patterns challenge the notion of nonlinearity as a universal characteristic of cognitive aging and suggest that such effects may be context-specific (Patterson 2018). This suggests that widowhood may act as an independent risk factor for cognitive decline, rather than amplifying age-related deterioration. The only exception was observed in ELSA, where a significant age × widowhood interaction in executive function was detected. Overall, the results suggest that the intersection of age and widowhood is not systematically associated with cognitive aging across cohorts. However, the lack of consistent nonlinear patterns or interaction effects should be interpreted cautiously because of the limits of sample size in some cohorts. Future research should further explore how age interacts with widowhood across different sociocultural and institutional environments.

Within the framework of established modifiable risk factors for dementia (Livingston et al. 2024), our findings offer additional insights into how individual characteristics relate to cognitive function among widowed older adults. Notably, educational attainment remained a robust protective factor for cognitive function (Zhang et al. 2024). The current findings suggest that internet use can decrease the likelihood of cognitive decline. Internet access or smartphone use, a low-cost way, can effectively promote social connections and can serve as a health management platform (Lu et al. 2022, James et al. 2025). Moreover, access to cognitive training apps and virtual reality may further reduce cognitive decline by providing targeted neuroplasticity exercises and immersive cognitive stimulation. A meta-analysis of 23 RCTs (N=894 stroke patients) found that VR-based therapies significantly improved executive function, memory, and visuospatial skills in patients with stroke (Zhang et al. 2021). In older populations and resource-limited settings, higher BMI may be a factor that can reduce the risk of cognitive decline among participants who lose a spouse (Lv et al. 2022). However, this interpretation requires caution, as reverse causation or survival bias may play a role. Further observational studies or clinical trials are needed to confirm our findings, as well as to develop tailored interventions to improve the widowed populations’ physical and mental health.

Strengths of this study include an extensive sample derived from nationally representative aging studies in high- and middle-income countries. This broad representation enhances the generalizability of the cross-cultural findings, facilitating meaningful comparisons between countries at varying levels of economic development. To better assess the cognitive performance of the elderly in diverse cultural backgrounds, we adopted standardized HCAP scores to assess cognitive function. Comprehensive statistical approaches—including generalized linear models, random-effects meta-analysis, and restricted cubic spines regressions—allowed us to capture both average effects and nonlinear trends in cognitive changes. Moreover, analyses restricted to widowed individuals offered insights into how individual factors may shape the effects of widowhood. Additionally, we conducted multiple imputations for the missing values and, after performing a series of sensitivity analyses, our results remained robust.

Nevertheless, certain limitations should be noted. First, given the significant increase in mortality risk following spousal loss (Wang et al. 2024), this may introduce selection bias, whereby only healthier individuals who survive are included in the study. Relatedly, reverse causation cannot be fully excluded, as pre-clinical cognitive decline may influence survival, participation, or marital trajectories. Second, we failed to identify the time-varying effects of being widowed (Hanes & Clouston 2021) and precise timing of spousal loss (Chen et al. 2022). Additionally, several factors relevant to widowhood research—including pre-loss marital quality (Min & Song 2023), caregiver status (Pinquart & Sörensen 2003), perceived social support, and cause of spousal death (Wu-Chung et al. 2025)—were not consistently measured in six cohorts and could not be incorporated into the harmonized analyses; future studies with richer psychosocial and environmental data are needed. Third, the exposure definition did not encompass other relationship statuses such as divorce, remarriage, or cohabitation. This restriction may introduce selection bias and limits generalizability beyond these groups. Future studies with more inclusive relationship measures are needed to extend these findings. Fourth, the minimum age of HCAP ranged from 50 years to 65 years, which might have led to the observed differences in associations, as age is a key determinant of cognitive performance. Finally, considering the cross-cultural nature of this study, we endeavored to define the variables in accordance with common protocols. However, significant heterogeneity was still observed in the pooled results, though our parallel analysis approach mitigated this concern.

## 5. Conclusions

Drawing on harmonized data from six longitudinal cohorts, this multinational study demonstrates that widowhood-associated cognitive changes exhibit domain-specific and sex-dependent patterns in diverse regions and socioeconomic contexts, with executive function and memory potentially emerging as the most consistently affected cognitive domain. And our findings illustrate how cognitive outcomes among widowed individuals are associated with individual and contextual factors, such as education, internet use, and BMI. These results highlight the importance of identifying protective mechanisms and tailoring public health strategies to vulnerable subgroups. Future studies should integrate biological and psychosocial measures to clarify mechanisms and inform targeted, context-sensitive interventions supporting cognitive resilience among widowed older adults, particularly in resource-limited settings.

## Data Availability

Original data from the HRS, ELSA, MHAS, CHARLS, LASI, and HAALSI are publicly available. The data can also be obtained on request.

## Ethical standards

HRS was approved by the Health Sciences and Behavioral Sciences Institutional Review Board (IRB-HSBS) at the University of Michigan. ELSA was approved by NHS Research Ethics Committees under the National Research and Ethics Service (NRES). MHAS was approved by the Institutional Review Boards of the University of Texas Medical Branch (UTMB) in the United States, the Instituto Nacional de Estadística y Geografía (INEGI) in Mexico), and the Instituto Nacional de Salud Pública (INSP) in Mexico. CHARLS was approved by the Institutional Review Board at Peking University. LASI obtained ethical approvals from the following collaborating organizations: Indian Council of Medical Research (ICMR), Delhi; IRB, International Institute for Population Sciences (IIPS), Mumbai; IRB, Harvard T.H. Chan School of Public Health (HSPH), Boston; IRB, University of Southern California (USC), Los Angeles; IRB, ICMRNational AIDS Research Institute (NARI), Pune; and IRB, Regional Geriatric Centres (RGCs), MoHFW. HAALSI was approved by the University of the Witwatersrand Human Research Ethics Committee, the Harvard T.H. Chan School of Public Health, Office of Human Research Administration and the Mpumalanga Provincial Research and Ethics Committee. All participants provided written informed consent. The authors assert that all procedures contributing to this work comply with the ethical standards of the relevant national and institutional committees on human experimentation and with the Helsinki Declaration of 1975, as revised in 2013.

## Acknowledgements.

We gratefully thank the administrations and data collection teams of the HRS, ELSA, MHAS, CHARLS, LASI, and HAALSI for providing publicly accessible datasets.

## Author Contributions

Conceptualization: Cancan Guo, Yue Wang and Fenfen Ge. Data curation: Cancan Guo. Formal analysis: Cancan Guo with the help of Yue Wang. Funding acquisition: Xinyu Sun. Supervision: Xinyu Sun and Fenfen Ge. Visualization: Yue Wang and Cancan Guo. Writing-original draft: Cancan Guo; Writing-review & editing: Fenfen Ge and Yue Wang

---

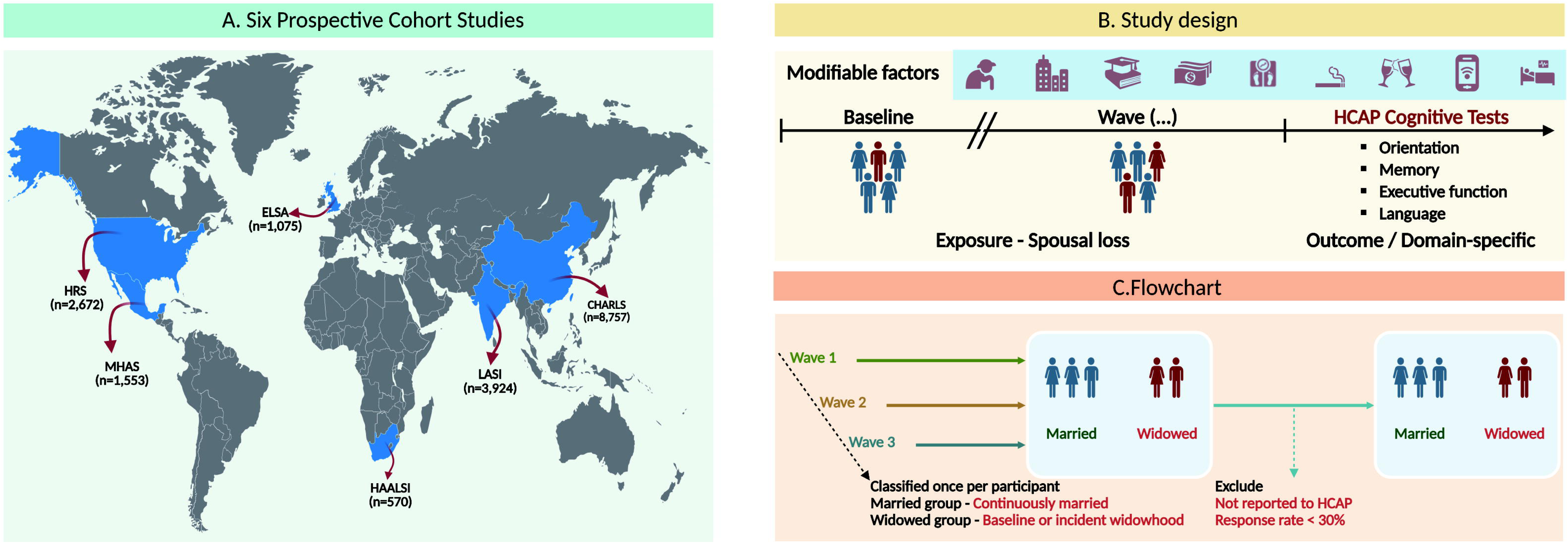

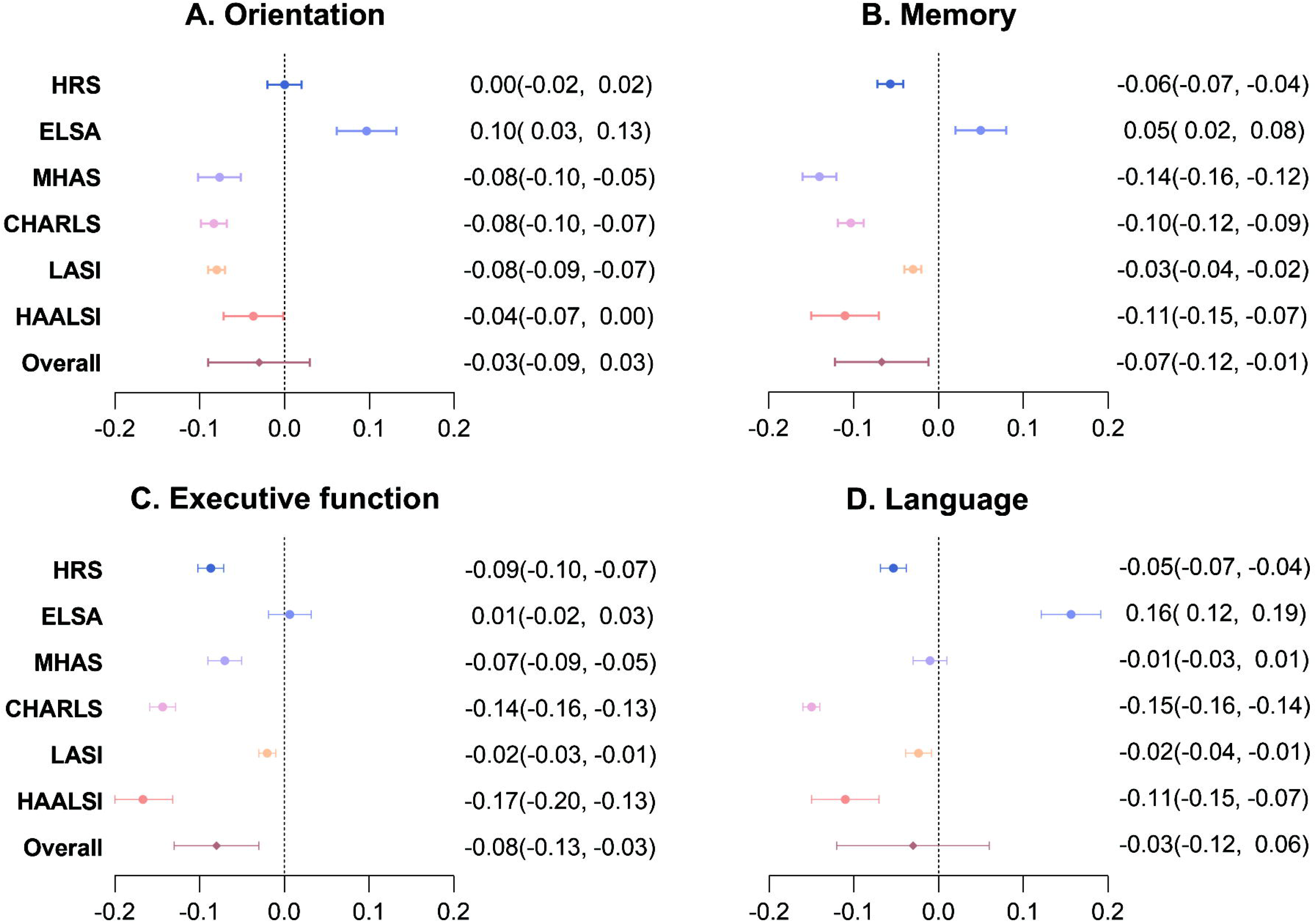

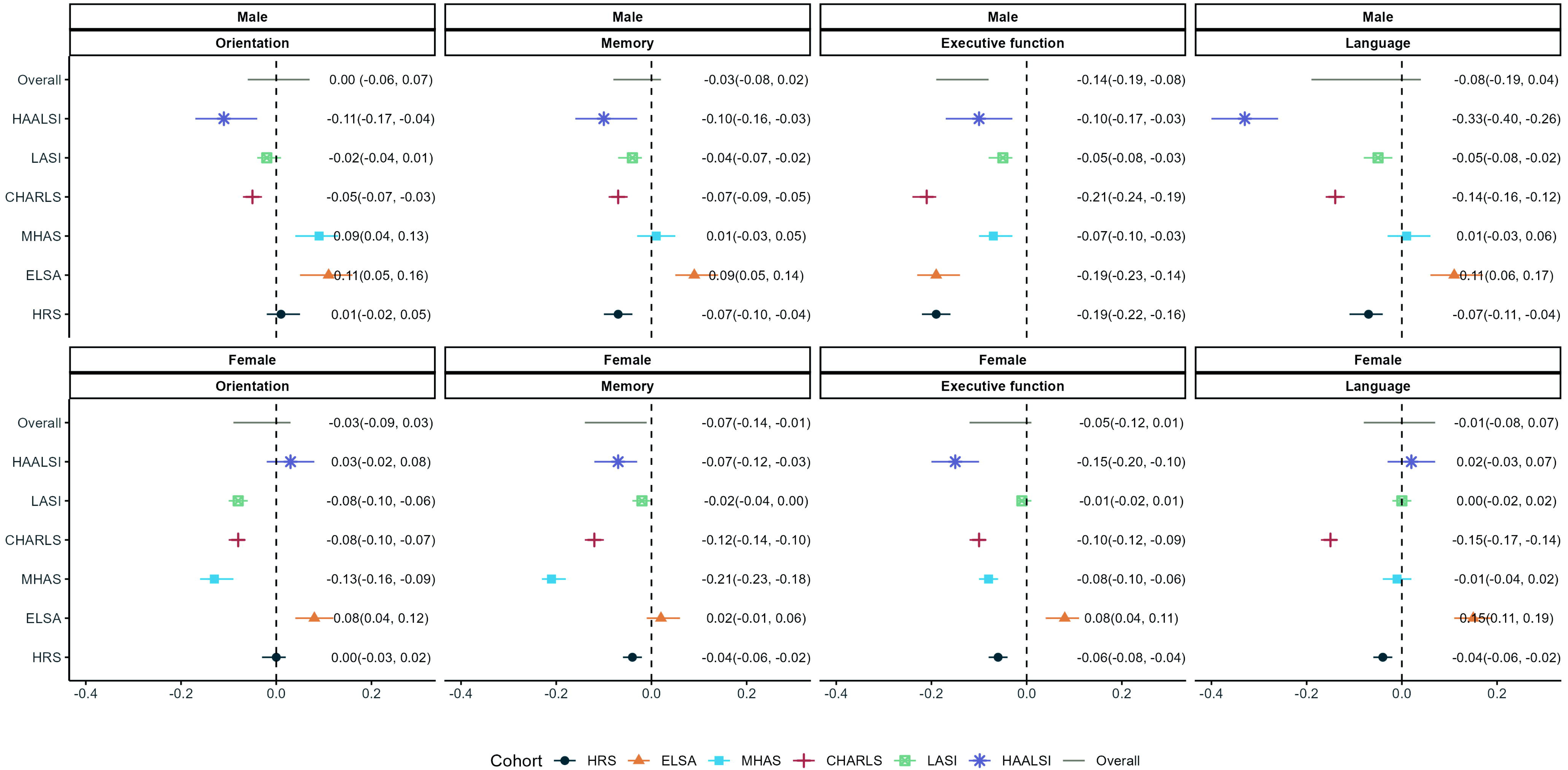

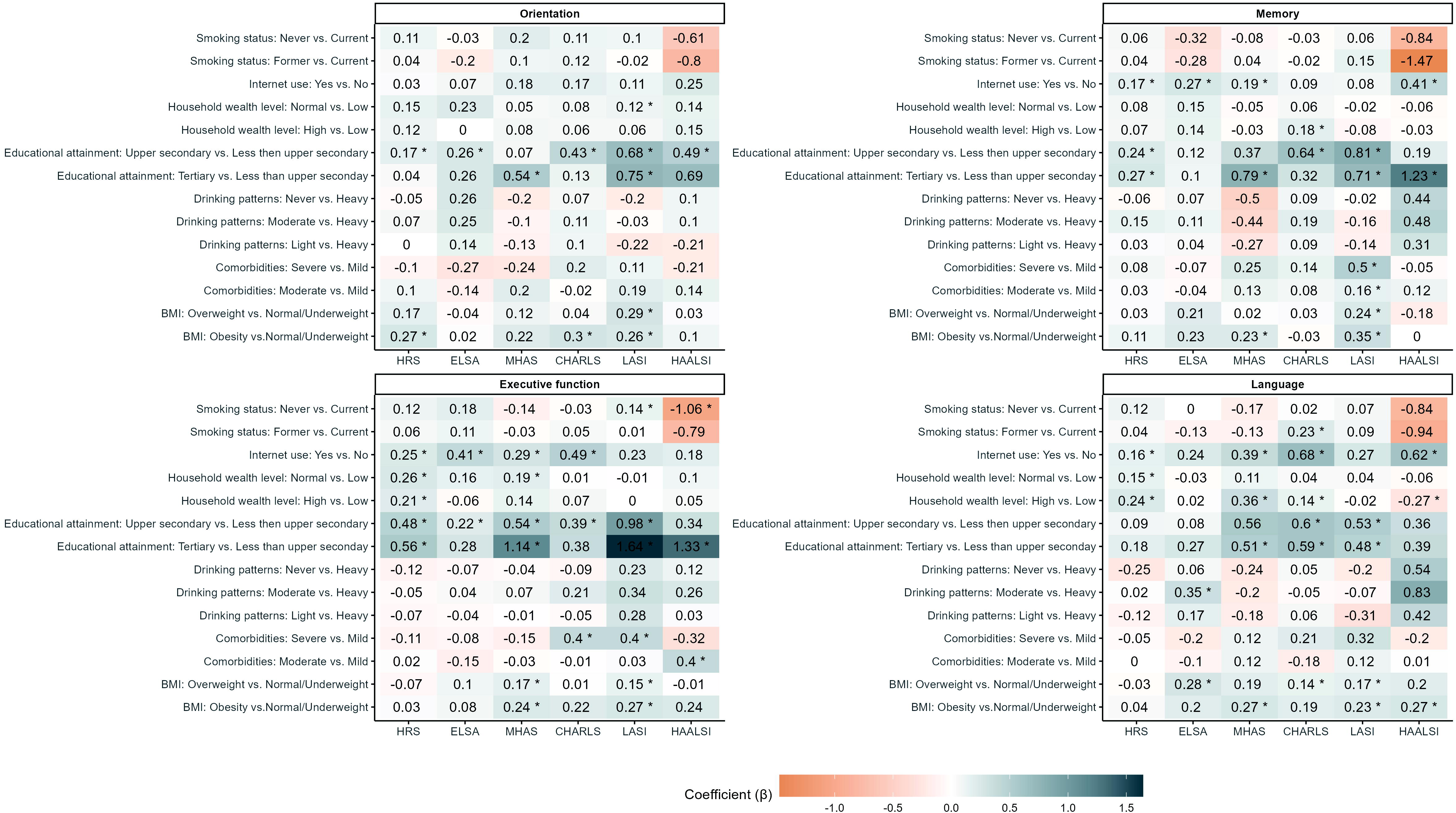

