## Supplementary Material for "Loss of a Spouse and Risk of Cognitive Decline: Insights from Six Prospective Cohort Studies"

Content

*[i. The Health and Retirement Study (HRS) and HRS-HCAP](#_Toc1548)*	[1](#_Toc1548)

*[ii. The English Longitudinal Study of Ageing (ELSA) and ELSA-HCAP](#_Toc4094)*	[1](#_Toc4094)

*[iii. The Mexican Health and Aging Study (MHAS) and Mex-cog](#_Toc3510)*	[2](#_Toc3510)

*[iv. The China Health and Retirement Longitudinal Study (CHARLS) and CHARLS-HCAP](#_Toc22488)*	[2](#_Toc22488)

*[v. The Longitudinal Aging Study in India (LASI) and LASI-DAD](#_Toc11716)*	[2](#_Toc11716)

*[vi. The Health and Aging in Africa: A Longitudinal Study of an INDEPTH Community in South Africa (HAALSI) and HAALSI-HCAP](#_Toc21625)*	[3](#_Toc21625)

*[Table S1: Harmonized definition of marital status in six aging studies](#_Toc24133)*	[4](#_Toc24133)

*[Table S2: Cognitive test items selected in each HCAP study in six aging studies](#_Toc2617)*	[5](#_Toc2617)

*[Table S3: Harmonized definition of covariates in six aging studies](#_Toc13860)*	[8](#_Toc13860)

*[Figure S1: Flow chart of participants selected in the HRS](#_Toc5313)*	[10](#_Toc5313)

*[Figure S2: Flow chart of participants selected in the ELSA](#_Toc7790)*	[10](#_Toc7790)

*[Figure S3: Flow chart of participants selected in the MHAS](#_Toc3273)*	[10](#_Toc3273)

*[Figure S4: Flow chart of participants selected in the CHARLS](#_Toc3400)*	[11](#_Toc3400)

*[Figure S5: Flow chart of participants selected in the LASI](#_Toc8200)*	[11](#_Toc8200)

*[Figure S6: Flow chart of participants selected in the HAALSI](#_Toc1600)*	[11](#_Toc1600)

*[Table S4: Characteristics of HRS participants by sex](#_Toc24730)*	[13](#_Toc24730)

*[Table S5: Characteristics of ELSA participants by sex](#_Toc20287)*	[14](#_Toc20287)

*[Table S6: Characteristics of MHAS participants by sex](#_Toc3525)*	[15](#_Toc3525)

*[Table S7: Characteristics of CHARLS participants by sex](#_Toc24192)*	[16](#_Toc24192)

*[Table S8: Characteristics of LASI participants by sex](#_Toc1120)*	[17](#_Toc1120)

*[Table S9: Characteristics of HAALSI participants by sex](#_Toc21438)*	[18](#_Toc21438)

*[Table S10: Overall meta-analysis estimates, 95% confidence interval, and I](#_Toc7002)^[2](#_Toc7002)^ [heterogeneity of specific-domain](#_Toc7002)*	[19](#_Toc7002)

*[Table S11: Male meta-analysis estimates, 95% confidence interval, and I](#_Toc23441)^[2](#_Toc23441)^ [heterogeneity of specific-domain](#_Toc23441)*	[19](#_Toc23441)

*[Table S12: Female meta-analysis estimates, 95% confidence interval, and I](#_Toc19494)^[2](#_Toc19494)^ [heterogeneity of specific-domain](#_Toc19494)*	[19](#_Toc19494)

*[Figure S7: Association between age and cognitive function by widowhood status in HRS, MHAS, and CHARLS](#_Toc22169)*	[20](#_Toc22169)

*[Figure S8: Association between age and cognitive function by widowhood status in ELSA, LASI, and HAALSI](#_Toc29911)*	[21](#_Toc29911)

*[Table S13: Individual-characteristics with cognitive function in the HRS widowed participants](#_Toc21114)*	[22](#_Toc21114)

*[Table S14: Individual-characteristics with cognitive function in the ELSA widowed participants](#_Toc4141)*	[24](#_Toc4141)

*[Table S15: Individual-characteristics with cognitive function in the MHAS widowed participants](#_Toc17582)*	[26](#_Toc17582)

*[Table S16: Individual-characteristics with cognitive function in the CHARLS widowed participants](#_Toc5266)*	[28](#_Toc5266)

*[Table S17: Individual-characteristics with cognitive function in the LASI widowed participants](#_Toc31261)*	[30](#_Toc31261)

*[Table S18: Individual-characteristics with cognitive function in the HAALSI widowed participants](#_Toc12681)*	[32](#_Toc12681)

*[Table S19: Association of between spousal loss and cognitive function using complete-case design](#_Toc3928)*	[34](#_Toc3928)

*[Table S20: Association of between spousal loss and cognitive function using a design that includes the participants only from baseline wave](#_Toc31318)*	[35](#_Toc31318)

*[Table S21: Association of between spousal loss and cognitive function using a design that restricts the HCAP participants age ≥ 65 years](#_Toc12791)*	[36](#_Toc12791)

*[Table S22: Associations between spousal loss and subsequent domain-specific cognitive function (Excluding HRS and ELSA)](#_Toc9577)*	[37](#_Toc9577)

**Ⅰ. Supplementary information**

The Health and Retirement Study (HRS) is a nationally representative, longitudinal, multidisciplinary survey of adults aged 50 years and older in the United States, together with a family of international sister studies designed to facilitate comparative research on aging. These cohorts share coordinated survey instruments, longitudinal follow-up, and harmonized data structures, enabling cross-national analyses across diverse socioeconomic and institutional contexts. To further standardize cognitive assessment, the Harmonized Cognitive Assessment Protocol (HCAP) was developed by the HRS team in collaboration with its sister studies to provide a comparable framework for measuring cognitive function worldwide. In each HCAP design, all the test items were based on a set of well-established cognitive and neuropsychological assessments and carefully translated into the local language, and the consistency has been previously demonstrated^1^.

***i. The Health and Retirement Study (HRS) and HRS-HCAP***^2,3^

The HRS is a prominent longitudinal panel study established in 1992 to investigate the health, economic, and social factors associated with aging in the United States. Since its inception, the HRS has been conducted every two years, surveying a nationally representative sample of about 20,000 adults aged 51 or older in the United States. The study is funded by the National Institute on Aging (NIA) and the Social Security Administration (SSA).

The Harmonized Cognitive Assessment Protocol (HCAP) Project, a sub-study within the HRS from 2016, is designed to measure a range of key cognitive domains affected by cognitive aging (including attention, memory, executive function, language, and visuospatial function) and to allow harmonization and comparisons to other studies in the US and around the world. And it randomly selecting HRS sample members aged 65 or older. The final HRS HCAP sample included 3,496 study subjects, representing a 79% response rate.

In this study, we used data from the RAND HRS Longitudinal File 2020 on the Gateway to Global Aging Data website and 2016 Harmonized Cognitive Assessment Protocol (HCAP) on the HRS website.

The HRS and HRS-HCAP received ethnical approval from the Health Sciences and Behavioral Sciences Institutional Review Board (IRB-HSBS) at the University of Michigan (HUM00061128).

***ii. The English Longitudinal Study of Ageing (ELSA) and ELSA-HCAP***^4,5^

The ELSA is a panel study of a representative cohort of men and women living in England aged 50 years. The study commenced in 2002, and the sample has been followed up every 2 years. The original sample consisted of 11 391 members ranging in age from 50 to 100 years. Funding for the ELSA is provided by the National Institute on Aging (NIA), the Office for National Statistics in Britain, and the British Heart Foundation.

The ELSA-HCAP involves an extensive range of cognitive measures on a stratified sub-sample of ELSA participants aged 65 and older. Participants were selected for the ELSA-HCAP Sub-study if they were an ELSA core member aged 65 and over at the start of fieldwork in January 2018 (born before 1 January 1953) and had completed an ELSA interview in person at either wave 8 (2016–17) or wave 7 (2014–15). A total of 1050 informant interviews were conducted, representing a response rate of 82.5% of all the eligible sample (n = 1273) contacted.

In this study, we used data from the Harmonized ELSA on the Gateway to Global Aging Data website and ELSA-HCAP on the UK Data Service website.

Ethical approval for all the ELSA waves was granted from NHS Research Ethics Committees under the National Research and Ethics Service (NRES).

***iii. The Mexican Health and Aging Study (MHAS) and Mex-cog***^6,7^

The MHAS was designed to examine the ageing process and its disease and disability burden of adults over the age of 50 in Mexico, with a highly comparable study protocols and survey instruments to the HRS. The first two waves were conducted in 2001 and 2003. And the third wave was conducted in 2012, with an every-three-year follow-up. The MHAS is partly sponsored by the National Institute on Aging (NIA) in the United States and the Instituto Nacional de Estadística y Geografía (INEGI) in Mexico.

The Cognitive Aging Ancillary Study in Mexico (Mex-Cog) performs an in-depth cognitive assessment in a subsample of older adults of the ongoing MHAS (Mex-Cog 2016 and 2021). A total of 2,265 subjects aged 55–104 years participated in 2016, representing a 70% response rate.

In this study, we used data from the Harmonized MHAS on the Gateway to Global Aging Data website and Mex-Cog 2016 on the MHAS website.

The study was approved by the Institutional Review Boards of the University of Texas Medical Branch (UTMB) in the United States, the Instituto Nacional de Estadística y Geografía (INEGI) in Mexico), and the Instituto Nacional de Salud Pública (INSP) in Mexico.

***iv. The China Health and Retirement Longitudinal Study (CHARLS) and CHARLS-HCAP***^8,9^

The CHARLS is a nationally representative longitudinal survey of adults aged 45 or older, examining health and economic adjustments to rapid ageing of the population in China. The CHARLS was established in 2011 and has been followed every 2 years, harmonized with leading international research studies in the HRS model. This work was funded by the National Institute of Health (NIH), the Natural Science Foundation of China (NSFC), the World Bank, and Peking University.

HCAP was added in wave 4 of CHARLS for respondents aged 60 and over to measure dementia. In Wave 4, some 11,021 respondents aged 60 and older, and their informants were part of the wave 4 CHARLS HCAP. The response rate was 99% out of all wave 4 respondents aged 60 and over.

In this study, we used data from the Harmonized CHARLS on the Gateway to Global Aging Data website and 2018 CHARLS Wave 4 on the CHARLS website.

Ethical approval for all the CHARLS waves was granted from the Institutional Review Board at Peking University (anthropometrics: IRB00001052-11015; biomarker collection: IRB00001052-11014).

***v. The Longitudinal Aging Study in India (LASI) and LASI-DAD***^10,11^

The LASI is a nationwide panel survey of adults aged 45 and older, collecting data on their health, social and economic well-being to understand aging and retirement in India. The study was established in 2016 and planned every 2–3 years for the next 25 years. The LASI is funded by the National Institute on Aging/National Institutes of Health (NIA/NIH) and the United Nations Population Fund–India office.

LASI-Diagnostic Assessment of Dementia (LASI-DAD) is an in-depth study of late-life cognition and dementia, aiming to better understand the determinants of late-life cognition, cognitive aging, and dementia in India. The LASI-DAD randomly drew a subsample of over 3,000 LASI respondents aged 60 and older, with a response rate reaching 82.9%.

In this study, we used data from the Harmonized LASI and LASI-DAD on the Gateway to Global Aging Data website.

Ethical approvals were obtained from the following collaborating organizations: Indian Council of Medical Research (ICMR), Delhi; IRB, International Institute for Population Sciences (IIPS), Mumbai; IRB, Harvard T.H. Chan School of Public Health (HSPH), Boston; IRB, University of Southern California (USC), Los Angeles; IRB, ICMRNational AIDS Research Institute (NARI), Pune; and IRB, Regional Geriatric Centres (RGCs), MoHFW.

***vi. The Health and Aging in Africa: A Longitudinal Study of an INDEPTH Community in South Africa (HAALSI) and HAALSI-HCAP***^12,13^

The HAALSI, a harmonized sister study to the HRS, was established to establish a population-based longitudinal cohort of men and women (2345 men and 2714 women) aged 40 and over in a rural South African community. It started in 2014 with longitudinal follow-up at 3-year intervals. The HAALSI is funded by the National Institute on Aging (NIA), and the Agincourt Health and Demographic Surveillance System site, the University of the Witwatersrand and Medical Research Council, South Africa, and the Wellcome Trust, UK.

The first wave of HAALSI Dementia (HAALSI-HCAP) sample was randomly drawn from 4176 participants enrolled in the second wave (2018–19) of HAALSI. And 690 HAALSI participants were invited to take part in the study.

In this study, we used data from HAALSI and HAALSI Dementia W2 on the HARVARD Dataverse website.

The study received ethical approvals from the University of the Witwatersrand Human Research Ethics Committee, the Harvard T.H. Chan School of Public Health, Office of Human Research Administration and the Mpumalanga Provincial Research and Ethics Committee.

**Ⅱ. Definition of key variables**

***Table S1: Harmonized definition of marital status in six aging studies***

| **HRS** | **ELSA** | **MHAS** | **CHARLS** | **LASI** | **HAALSI** |
| --- | --- | --- | --- | --- | --- |
| 1. Married^a^  2. Married, spouse absent  4. Separated  5. Divorced  6. Separated/divorced  7. Widowed^b^  8. Never married  9. Unknown unmar | 1. Married^a^  3. Partnered  4. Separated  5. Divorced  7. Widowed^b^  8. Never married | 1. Married^a^  3. Partnered  4. Separated  5. Divorced  7. Widowed^b^  8. Never married | 1. Married^a^  3. Partnered  4. Separated  5. Divorced  7. Widowed^b^  8. Never married | 1. Married^a^  3. Partnered  4. Separated  5. Divorced  7. Widowed^b^  8. Never married | 1. Never married  2. Currently married or living with partner^a^  3. Separated/Deserted  4. Divorced  5. Widowed^b^ |

*Note: a for married status; b for spousal loss*

*HRS, the Health and Retirement Study; ELSA, the English Longitudinal Study of Ageing; MHAS, the Mexican Health and Aging Study; CHARLS, the China Health Retirement Longitudinal Study; LASI, the Longitudinal Aging Study in India; HAALSI, the Health and Aging in Africa: A Longitudinal Study of an INDEPTH Community in South Africa.*

***Table S2: Cognitive test items selected in each HCAP study in six aging studies***

We used standardized *z* scores of domain-specific cognitive functions (orientation, memory, executive function, and language) as the primary outcome in this study. Additionally, due to item response rate being below 80%, we removed several cognitive test items from the ELSA, CHARLS, LASI, and HAALSI to minimize statistical bias by reducing the impact of missing data.

| **Variables** | **HRS** | **ELSA** | **MHAS** | **CHARLS** | **LASI** | **HAALSI** |
| --- | --- | --- | --- | --- | --- | --- |
| **Orientation** | | | | | | |
| Date | √ | √ | √ | √ | √ | √ |
| Month | √ | √ | √ | √ | √ | √ |
| Year | √ | √ | √ | √ | √ | √ |
| Day of week | √ | √ | √ | √ | √ | √ |
| Season of year | √ | √ | - | √ | √ | √ |
| What time is it? | - | - | √ | - | - | √ |
| Country | - | √ | √ | - | - | √ |
| State | √ | - | √ | √ | √ | √ |
| County | √ | √ | - | √ | - | √ |
| City or town | √ | √ | - | √ | √ | - |
| Address of building | √ | √ | √ | √ | √ | √ |
| Name of building | - | √ | - | - | √ | - |
| Floor of building | √ | - | - | √ | √ | √ |
| **Memory** | | | | | | |
| CERAD immediate sum of 3 trials | √ | √ | √ | √ | √ | √ |
| CERAD word list delay | √ | √ | √ | √ | √ | √ |
| CERAD recognition | √ | √ | √ | √ | √ | √ |
| Three-word immediate registration | √ | √ | √ | √ | √ | √ |
| Three-word delayed recall | √ | √ | √ | √ | √ | √ |
| Logical Memory immediate | √ | √ | √ | - | √ | √ |
| Logical Memory delay | √ | √ | √ | - | √ | √ |
| Logical memory recognition | √ | √ | - | - | √ | √ |
| Brave man immediate (East Boston Memory Test) | √ | √ | √ | - | √ | - |
| Brave man delay (East Boston Memory Test) | √ | √ | √ | - | √ | - |
| CERAD constructional praxis delay | √ | √ | √ | - | √ | × |
| **Executive function** | | | | | | |
| Backward counting, 20-0 | - | - | √ | - | - | - |
| Backward counting, 100-0 | √ | √ | - | - | - | - |
| Forward day naming | - | - | - | - | - | √ |
| Backward Day naming | - | - | - | - | √ | √ |
| CDR calculation-cent | - | - | - | - | - | √ |
| Digit Span Forward (single item) | - | - | - | - | √ | - |
| Digit Span Backward (single item) | - | - | - | - | √ | - |
| Digit Span Forward (multiple items) | - | - | - | - | - | √ |
| Digit Span Backward (multiple items) | - | - | - | - | - | √ |
| Go-No-Go | - | - | √ | - | √ | √ |
| Letter cancellation | √ | √ | - | - | - | - |
| Motor Programming | - | - | - | - | - | √ |
| Number series | √ | √ | - | × | - | - |
| Problem solving | - | - | - | - | × | - |
| Raven’s progressive matrices | √ | √ | - | - | √ | √ |
| Serial 3s | - | - | √ | - | - | - |
| Serial 7s | - | √ | √ | √ | × | × |
| Similarities | - | - | √ | - | × | √ |
| Spelling backwards | √ | × | - | - | - | × |
| Symbols and Digits test | √ | √ | √ | - | - | - |
| Symbol Cancellation Test | - | - | √ | - | √ | √ |
| Token Test | - | - | - | - | × | √ |
| Trails A time (letters and numbers) | √ | √ | - | - | - | - |
| Trails B time (letters and numbers) | √ | √ | - | - | - | - |
| **Language** | | | | | | |
| Animal fluency | √ | √ | √ | √ | √ | √ |
| Name a described cactus | √ | √ | - | √ | - | √ |
| Name a described coconut | - | - | - | - | √ | - |
| Name the elbow | √ | √ | √ | √ | √ | √ |
| Name president or Prime Minister | √ | √ | - | √ | √ | √ |
| Name deputy president | - | - | - | - | - | √ |
| Object naming (watch) | √ | √ | - | √ | √ | √ |
| Object naming (pencil) | √ | √ | √ | √ | √ | √ |
| Object naming (shoe) | - | - | √ | - | - | - |
| Define Bridge | - | - | √ | - | - | - |
| What are scissors used for? | √ | √ | √ | √ | √ | √ |
| What does one do with a hammer? | √ | √ | √ | √ | √ | √ |
| Where is the local market? | √ | √ | √ | √ | √ | √ |
| Point to 2 things in the vicinity | √ | √ | √ | √ | √ | √ |
| Write or say a sentence | √ | √ | √ | × | √ | × |
| Repetition of a phrase | √ | √ | √ | √ | √ | √ |
| Read and follow command | √ | √ | √ | √ | √ | × |
| Follow 3-stage instruction | √ | √ | √ | √ | × | √ |
| Boston Naming Test | - | - | - | - | - | √ |

*Note: √ for those items that are consisted in respective HCAP and are applied in this study; × for those items that are consisted in respective HCAP but are not applied in this study because the response rate below 80%; - for those items that are not administrated in retrospective HCAP.*

*HRS, the Health and Retirement Study; ELSA, the English Longitudinal Study of Ageing; MHAS, the Mexican Health and Aging Study; CHARLS, the China Health Retirement Longitudinal Study; LASI, the Longitudinal Aging Study in India; HAALSI, the Health and Aging in Africa: A Longitudinal Study of an INDEPTH Community in South Africa.*

***Table S3: Harmonized definition of covariates in six aging studies***

| **Covariates** | **HRS** | **ELSA** | **MHAS** | **CHARLS** | **LASI** | **HAALSI** |
| --- | --- | --- | --- | --- | --- | --- |
| Age (in years) | It is the integer portion of the number of months old divided by 12. And the study age criteria are not less than 50 years. | It is provided directly by ELSA in the Core files and top-coded at 90 years old, which means all respondents who report an age of 90 or older have a value of 90 years. And the study age criteria are not less than 50 years. | It comes from the recorded age in the sampling directory and is also calculated using the self-reported birth year and the year of interview because of missing sampling directory data. And the study age criteria are not less than 50 years. | It is calculated using the interview year and month and the respondent's birth year and month. And the study age criteria are not less than 45 years. | It is the cleaned current age in years at the time of the current wave’s interview from self-reported birth year, self-reported current age, and current age as reported in the household roster. And the study age criteria are not less than 45 years. | It comes from census at the time of sample selection on July 31, 2014. And the study age criteria are not less than 40 years. |
| Sex | male/female | | | | | |
| Minority status^14^ | In different countries, the definition of minority group varies based on historical, cultural, and socio-economic contexts. In the United States and England, minority group is typically defined by race or ethnicity, reflecting disparities in income, education, and health^15,16^. In contrast, in developing countries like China, Mexico, South Africa, and India, rural populations are often considered minority group due to stark urban-rural disparities in resources, healthcare, and education^17^. Rural residents face lower life expectancy and poorer health status, making them an important focus in health and social research. | | | | | |
|  | Race is assigned by looking at reports from Tracker and all waves of data, which is divided into White/Caucasian and Non-white (Black/African American and other races). In this study, the non-white populations belong to minority group. | Race is assigned by looking at reports from all waves of data, which is divided into White and Non-white. In this study, the non-white populations belong to minority group. | It indicates whether the respondent's household resides in an urban or rural location. Participants who live in rural village are recorded as minority group. | It indicates the household living region (urban community or rural village), which is defined by National Bureau of Statistics of the People's Republic of China. Participants who live in rural village are recorded as minority group. | It indicates the respondent's living region (urban community or rural village). Participants who live in rural village are recorded as minority group. | The HAALSI study did not include minority status group as covariates, as it exclusively sampled from Black Africans population in rural areas (Agincourt, South Africa)^12^. |
| Educational attainment | It identifies the highest level of education completed according to a three-tier harmonized scale (less than upper secondary, upper secondary and vocational training, tertiary education) from a simplified version of 1997 International Standard Classification of Education (ISCED-97) which was developed to compare education levels across countries^18^. | | | | | |
|  | 1.less than upper secondary education: less than high school.  2.upper secondary and vocational training: GED, high-school graduate, and some college.  3.tertiary education: college and above. | 1.less than upper secondary education: no qualification.  2.upper secondary and vocational training: nvq1/cse other grade equiv, and nvq2/gce o level equiv.  3.tertiary education: nvq3/gce a level equiv, nvq4/nvq5/degree or equiv, and higher ed below degree. | 1.less than upper secondary education: no formal education, primary school  2.upper secondary and vocational training: preparatory and high school.  3.tertiary education: basic teaching school, college, and graduate school. | 1.less than upper secondary education: no formal education (illiterate), did not finish primary school but can read, Sishu (private tutoring), elementary school, and middle school.  2.upper secondary and vocational training: high school and vocational school. 3.tertiary education: two/three-year college, college grad, and post-graduate degree. | 1.less than upper secondary education: no formal education, less than primary school, primary school.  2.upper secondary and vocational training: middle school, secondary school/matriculation, higher secondary/intermediate/senior secondary, and diploma and certificate holders.  3.tertiary education: graduate degree, post-graduate degree and above, and professional course/degree. | 1.less than upper secondary education: no formal education, preschool, Sub-A/Gr 1, Sub-B/Gr 2, Std 1/Gr 3, Std 2/Gr 4, Std 3/Gr 5, Std 4/Gr 6, Std 5/Gr 7, Std 6/Gr 8, Std 7/Gr 9, and Std 8/Gr 9.  2.upper secondary and vocational training: Std 9/Gr 11, Stad 10/Gr 12/Matric, ABET 1, ABET 2, ABET 3, and ABET 4.  3.tertiary education: partial tertiary, completed tertiary, partial university, and university. |
| Household wealth level^19^ | We use household per capital consumption (PCE) within each country as a measure of household wealth level. The PCE in tertiles was divided into low, normal and high level of household wealth level. | | | | | |
| Body mass index, BMI | 1. BMI is a numerical measure calculated from a person's weight and height and calculated by using weight in kilograms (kg) divided by the square of height in meters (m^2^).  2. BMI types:  (1) underweight or normal weight: <25.0 kg/m^2^,  (2) overweight: 25.0–29.9 kg/m^2^,  (3) obesity: ≥30.0 kg/m^2^. | | | | | |
| Smoking status | It indicates the participants’ smoking status, specifically including never smoked, former smoker (having quit), and current smoker, which is obtained from the self-reported smoking history. | | | | | |
| Drinking patterns | It reflects the drinking patterns formed based on the self-reported drinking frequency (daily, weekly, or monthly)of the participants, specifically classified as never drinking, light drinking, moderate drinking and heavy drinking. | | | | | |
|  | 1. never drinking.  2. light drinking: current drinker but drink less than once a week.  3. moderate drinking: drink 1 to 4 days a week.  4. heavy drinking: drink at least 5 days a week. | 1. never drinking.  2. light drinking: current drinker but drink less than once a week.  3. moderate drinking: drink 1 to 4 days a week.  4. heavy drinking: drink at least 5 days a week. | 1. never drinking.  2. light drinking: current drinker but drink less than once a week.  3. moderate drinking: drink 1 to 4 days a week.  4. heavy drinking: drink at least 5 days a week. | 1. never drinking.  2. light drinking: current drinker but drink less than once a week.  3. moderate drinking: drink 1 to 3 days a week.  4. heavy drinking: drink at least 4 days a week. | 1. never drinking.  2. light drinking: current drinker but drink less than once a week.  3. moderate drinking: drink 1 to 4 days a week.  4. heavy drinking: drink at least 5 days a week. | 1. never drinking.  2. light drinking: current drinker but drink less than once a week.  3. moderate drinking: drink 1 to 4 days a week.  4. heavy drinking: drink at least 5 days a week. |
| Internet use^20^ | It indicates the participants’ access and capability to use Information and Communications Technologies (ICTs) such as the internet and can thus be classified as digital inclusion or exclusion according to the frequency of interacting with others using internet, phone, or email. | | | | | |
|  | Question: Do you regularly use the Internet (or the World Wide Web) for sending and receiving e-mail or for any other purpose, such as making purchases, searching for information, or making travel reservations?  The response “no” was defined as digital exclusion, while the response “yes” was recorded as digital inclusion. | Question: On average, how often do you use the Internet or email?  A frequency of less than once a week was defined as digital exclusion, while a frequency of at least once a week was recorded as digital inclusion. | Question: Have you talked on the phone with relatives or friends or used the computer to send messages or used the internet last week?  The response “no” was defined as digital exclusion, while the response “yes” was recorded as digital inclusion. | Question: Have you used the Internet in the past month?  The response “no” was defined as digital exclusion, while the response “yes” was recorded as digital inclusion. | Question: How often do you use a computer for email or net surfing?  A frequency of less than once a week was defined as digital exclusion, while a frequency of at least once a week was recorded as digital inclusion. | Question: How often did you typically interact with this person on the phone, by SMS, through email or the internet over the past 6 months?  A frequency of less than once a week was defined as digital exclusion, while a frequency of at least once a week was recorded as digital inclusion. |
| Comorbidities | In this study, we selected four non-communicable diseases (NCDs) to assess the severity of participants' comorbidity, including hypertension, diabetes, stroke and heart disease. According to the number of the above diseases that participants suffered from, the severity of comorbidity was classified into mild (0 to 1 disease), moderate (2 diseases), and severe (3 to 4 diseases). | | | | | |
| Baseline cognition^21^ | In this study, we used the quartiles of baseline episodic memory scores to characterize the participants’ baseline cognitive function. Episodic memory mainly includes immediate and delayed word recall performance. For HRS, CHARLS, HAALSI, ELSA and LASI, the total score is 20 points; while for MHAS, the total score is 16 points. | | | | | |

*HRS, the Health and Retirement Study; ELSA, the English Longitudinal Study of Ageing; MHAS, the Mexican Health and Aging Study; CHARLS, the China Health Retirement Longitudinal Study; LASI, the Longitudinal Aging Study in India; HAALSI, the Health and Aging in Africa: A Longitudinal Study of an INDEPTH Community in South Africa.*

**Ⅲ. Flow charts**

We conducted a longitudinal design with the exposure of spousal loss measured at parent studies, matching with cognitive outcomes from HCAP sub-studies. Participants who reported their marital status and fulfilled the age eligibility criteria within their countries were included; for instance, those aged ≥ 50 years in HRS and ≥ 65 years in HRS-HCAP. We excluded participants who had missing data exceeding 30% for the HCAP cognitive test or covariate items.

***Figure S1: Flow chart of participants selected in the HRS***

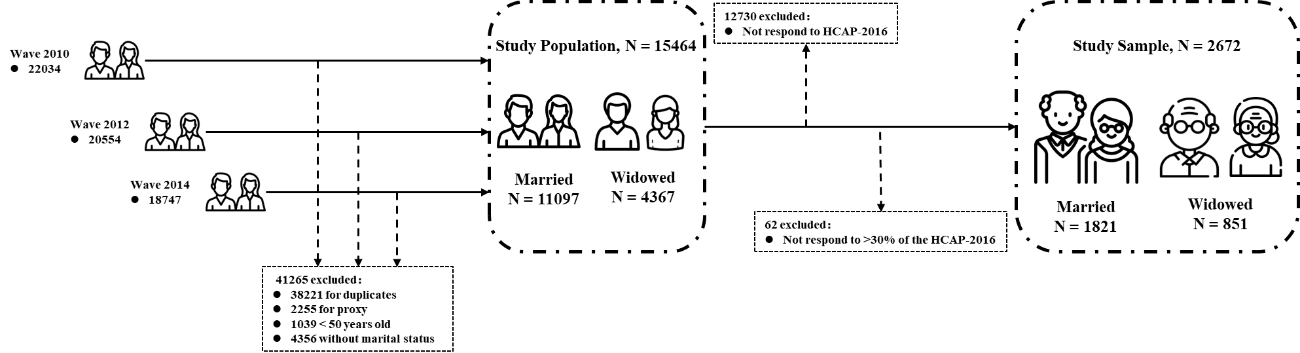

*HRS, the Health and Retirement Study.*

***Figure S2: Flow chart of participants selected in the ELSA***

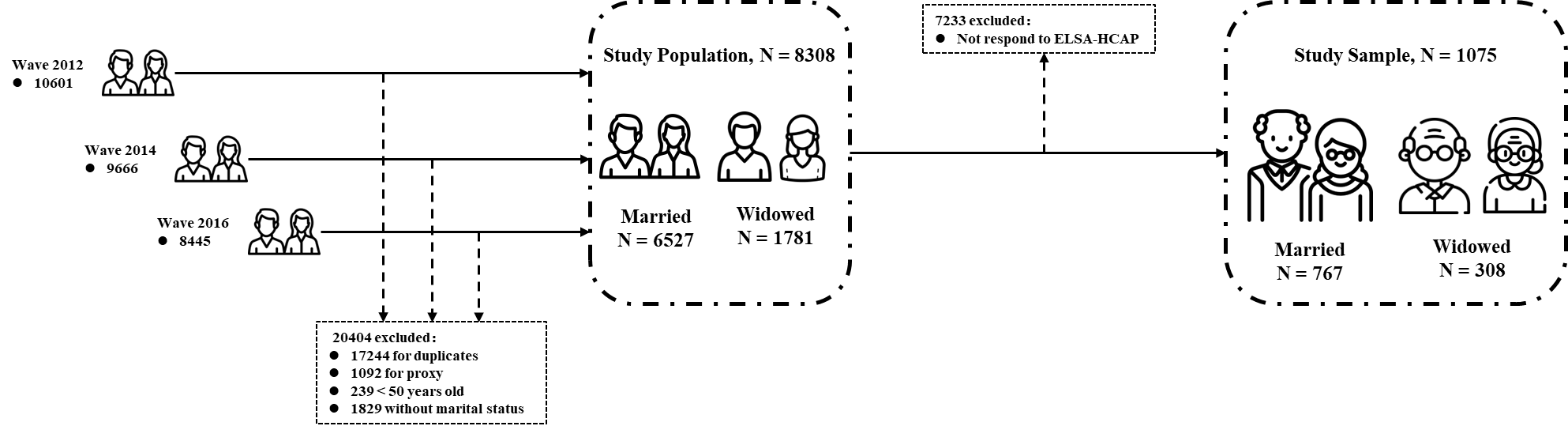

*ELSA, the English Longitudinal Study of Ageing.*

***Figure S3: Flow chart of participants selected in the MHAS***

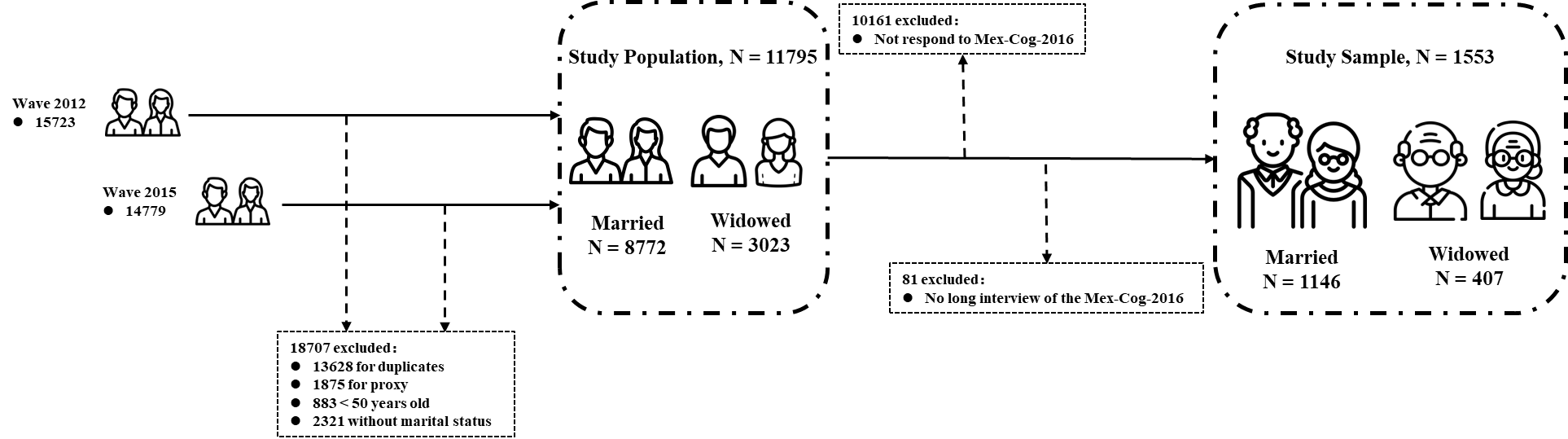

*MHAS, the Mexican Health and Aging Study.*

***Figure S4: Flow chart of participants selected in the CHARLS***

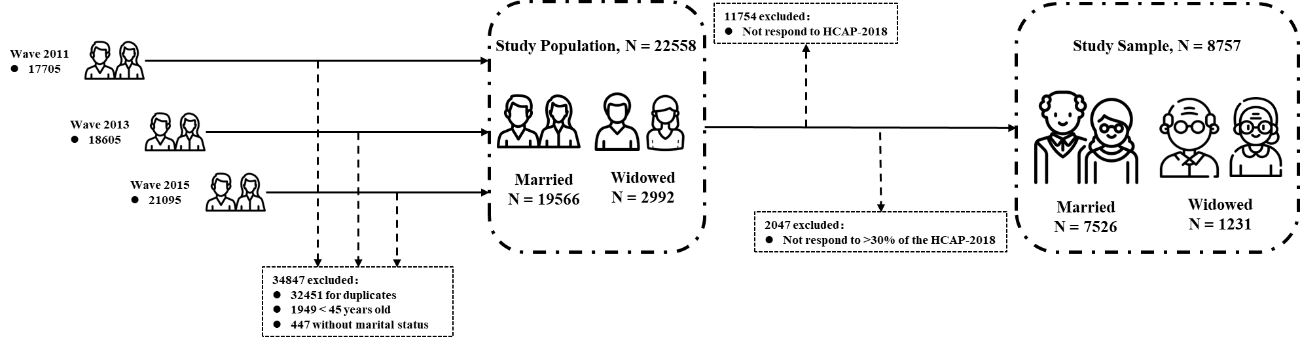

*CHARLS, the China Health Retirement Longitudinal Study.*

***Figure S5: Flow chart of participants selected in the LASI***

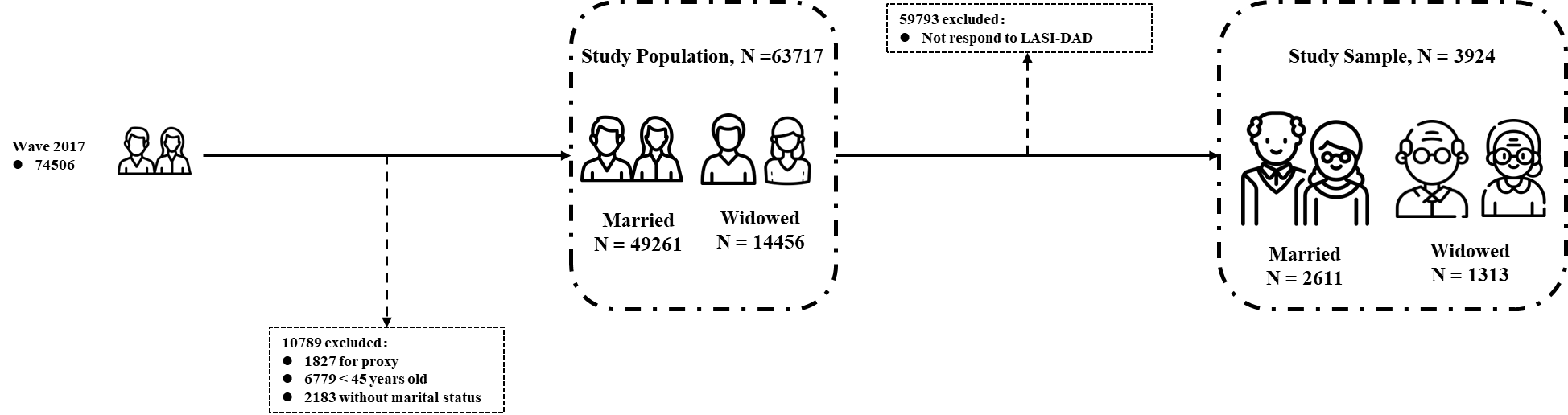

*LASI, the Longitudinal Aging Study in India.*

***Figure S6: Flow chart of participants selected in the HAALSI***

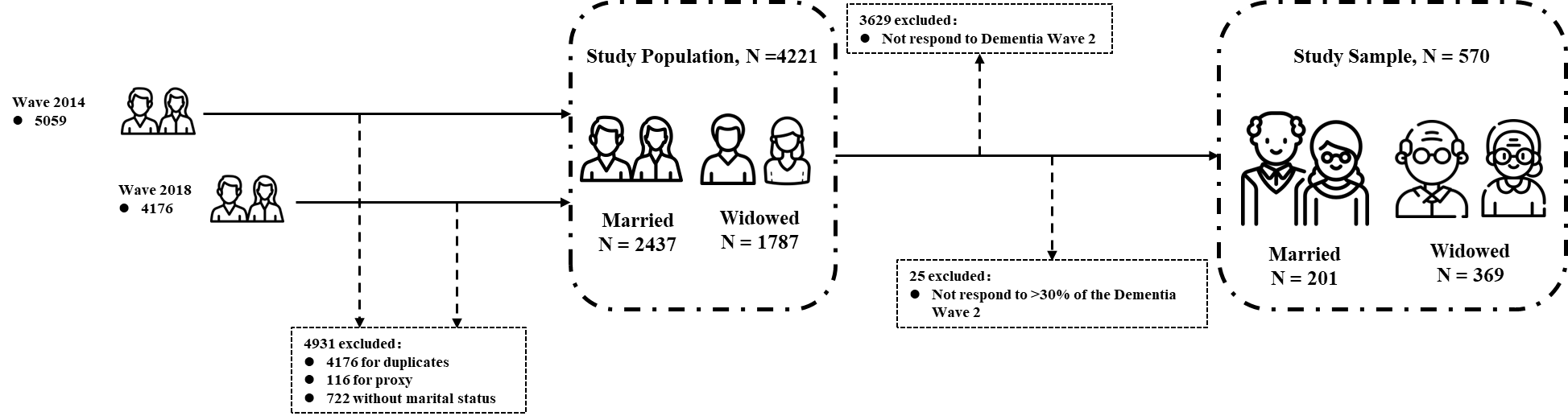

*HAALSI, the Health and Aging in Africa.*

**Ⅳ. Supplementary methods**

This study employed a parallel, cohort-specific analytic framework using harmonized data from six population-based aging cohorts. Analyses were conducted independently within each cohort using comparable model specifications, and effect estimates were subsequently synthesized using meta-analytic methods to assess consistency and heterogeneity across settings.

This approach aligns with methodological recommendations for cross-national research using the Harmonized Cognitive Assessment Protocol (HCAP), which emphasize the importance of accounting for contextual variation when examining socially embedded exposures across countries ^22^. Under this framework, parallel analyses enable estimation of cohort-specific associations while maintaining analytic comparability, without imposing assumptions of uniform effect structures across diverse cultural and socioeconomic contexts.

In the present study, spousal loss was conceptualized as a life-course exposure whose meaning, timing, and social consequences may vary across populations. The parallel analytic framework therefore facilitates transparent comparison of country-specific effect estimates and supports evaluation of cross-national heterogeneity through meta-analytic synthesis.

Additionally, outliers were identified by the quartile method and subsequently converted into missing values. We performed multiple imputation by chained equations to adequately address missing data in all analyses^23^.

**Ⅴ. Baseline characteristics of participants**

***Table S4: Characteristics of HRS participants by sex***

|  | | **Male(*n*=1077)** | | ***p* value** | **Female(*n*=1595)** | | ***p* value** |
| --- | --- | --- | --- | --- | --- | --- | --- |
|  |  | **Married**  ***n*=930 (86.3)** | **Widowed**  ***n*=147 (13.7)** |  | **Married**  ***n*=891 (55.9)** | **Widowed**  ***n*=704 (44.1)** |  |
| **Age (in years)** |  | 69.60 (7.02) | 74.93 (7.52) | <0.001 | 68.33 (6.61) | 73.78 (7.72) | <0.001 |
| **Minority status** | White/ Caucasian | 788 (84.7) | 120 (81.6) | 0.566 | 755 (84.7) | 533 (75.7) | <0.001 |
|  | Non-white | 141 (15.2) | 27 (18.4) |  | 136 (15.3) | 171 (24.3) |  |
|  | Missing | 1 (0.1) | 0 (0.0) |  | 0 (0.0) | 0 (0.0) |  |
| **Educational attainment** | Less than upper secondary | 127 (13.7) | 33 (22.4) | 0.003 | 131 (14.7) | 170 (24.1) | <0.001 |
|  | Upper secondary or vocational training | 502 (54.0) | 85 (57.8) |  | 526 (59.0) | 430 (61.1) |  |
|  | Tertiary | 300 (32.3) | 29 (19.7) |  | 234 (26.3) | 104 (14.8) |  |
|  | Missing | 1 (0.1) | 0 (0.0) |  | 0 (0.0) | 0 (0.0) |  |
| **Household wealth level** | Low | 253 (27.2) | 67 (45.6) | <0.001 | 201 (22.6) | 160 (22.7) | <0.001 |
|  | Normal | 316 (34.0) | 41 (27.9) |  | 270 (30.3) | 184 (26.1) |  |
|  | High | 361 (38.8) | 39 (26.5) |  | 359 (40.3) | 288 (40.9) |  |
|  | Missing | 0 (0.0) | 0 (0.0) |  | 0 (0.0) | 0 (0.0) |  |
| **BMI** | Underweight/Normal | 114 (12.3) | 20 (13.6) | 0.019 | 201 (22.6) | 160 (22.7) | 0.050 |
|  | Overweight | 360 (38.7) | 57 (38.8) |  | 270 (30.3) | 184 (26.1) |  |
|  | Obesity | 393 (42.3) | 50 (34.0) |  | 359 (40.3) | 288 (40.9) |  |
|  | Missing | 63 (6.8) | 20 (13.6) |  | 61 (6.8) | 72 (10.2) |  |
| **Smoking status** | Current | 85 (9.1) | 16 (10.9) | 0.266 | 61 (6.8) | 59 (8.4) | 0.599 |
|  | Former | 523 (56.2) | 89 (60.5) |  | 325 (36.5) | 254 (36.1) |  |
|  | Never | 317 (34.1) | 40 (27.2) |  | 500 (56.1) | 385 (54.7) |  |
|  | Missing | 5 (0.5) | 2 (1.4) |  | 5 (0.6) | 6 (0.9) |  |
| **Drinking patterns** | Heavy | 139 (14.9) | 26 (17.7) | 0.699 | 91 (10.2) | 43 (6.1) | <0.001 |
|  | Moderate | 296 (31.8) | 50 (34.0) |  | 215 (24.1) | 122 (17.3) |  |
|  | Light | 162 (17.4) | 24 (16.3) |  | 167 (18.7) | 115 (16.3) |  |
|  | Never | 333 (35.8) | 47 (32.0) |  | 418 (46.9) | 424 (60.2) |  |
|  | Missing | 0 (0.0) | 0 (0.0) |  | 0 (0.0) | 0 (0.0) |  |
| **Internet use** | No | 383 (41.2) | 93 (63.3) | <0.001 | 352 (39.5) | 460 (65.3) | <0.001 |
|  | Yes | 547 (58.8) | 54 (36.7) |  | 539 (60.5) | 244 (34.7) |  |
|  | Missing | 0 (0.0) | 0 (0.0) |  | 0 (0.0) | 0 (0.0) |  |
| **Comorbidities** | Mild | 607 (65.3) | 82 (55.8) | 0.067 | 658 (73.8) | 444 (63.1) | <0.001 |
|  | Moderate | 226 (24.3) | 48 (32.7) |  | 175 (19.6) | 190 (27.0) |  |
|  | Severe | 97 (10.4) | 17 (11.6) |  | 58 (6.5) | 70 (9.9) |  |
|  | Missing | 0 (0.0) | 0 (0.0) |  | 0 (0.0) | 0 (0.0) |  |
| **Baseline cognition** | ≤25% | 330 (35.5) | 78 (53.1) | 0.001 | 198 (22.2) | 250 (35.5) | <0.001 |
|  | 25~50% | 263 (28.3) | 33 (22.4) |  | 210 (23.6) | 179 (25.4) |  |
|  | 50%~75% | 180 (19.4) | 22 (15.0) |  | 236 (26.5) | 157 (22.3) |  |
|  | ＞75% | 157 (16.9) | 14 (9.5) |  | 247 (27.7) | 118 (16.8) |  |
|  | Missing | 6 (0.6) | 0 (0.0) |  | 0 (0.0) | 0 (0.0) |  |

*Note: Data are N (%) for categorical variables or mean (SD) for continuous variables.*

*HRS, the Health and Retirement Study; BMI, body mass index.*

***Table S5: Characteristics of ELSA participants by sex***

|  | | **Male (*n*=475)** | | ***p* value** | **Female (n=600)** | | ***p* value** |
| --- | --- | --- | --- | --- | --- | --- | --- |
|  |  | **Married**  ***n*=407 (85.7)** | **Widowed**  ***n*=68 (14.3)** |  | **Married**  ***n*=360 (60.0)** | **Widowed**  ***n*=240 (40.0)** |  |
| **Age (in years)** |  | 69.77 (6.58) | 76.07 (7.45) | <0.001 | 68.62 (5.98) | 75.50 (7.25) | <0.001 |
| **Minority status** | White/ Caucasian | 390 (95.8) | 65 (95.6) | 1.000 | 352 (97.8) | 237 (98.8) | 0.576 |
|  | Non-white | 17 (4.2) | 3 (4.4) |  | 8 (2.2) | 3 (1.2) |  |
|  | Missing | 0 (0.0) | 0 (0.0) |  | 0 (0.0) | 0 (0.0) |  |
| **Educational attainment** | Less than upper secondary | 99 (24.3) | 26 (38.2) | 0.006 | 124 (34.4) | 131 (54.6) | <0.001 |
|  | Upper secondary or vocational training | 219 (53.8) | 29 (42.6) |  | 153 (42.5) | 68 (28.3) |  |
|  | Tertiary | 76 (18.7) | 7 (10.3) |  | 34 (9.4) | 9 (3.8) |  |
|  | Missing | 13 (3.2) | 6 (8.8) |  | 49 (13.6) | 32 (13.3) |  |
| **Household wealth level** | Low | 90 (22.1) | 34 (50.0) | <0.001 | 74 (20.6) | 159 (66.2) | <0.001 |
|  | Normal | 147 (36.1) | 17 (25.0) |  | 137 (38.1) | 56 (23.3) |  |
|  | High | 170 (41.8) | 16 (23.5) |  | 149 (41.4) | 22 (9.2) |  |
|  | Missing | 0 (0.0) | 1 (1.5) |  | 0 (0.0) | 3 (1.2) |  |
| **BMI** | Underweight/Normal | 76 (18.7) | 15 (22.1) | 0.316 | 104 (28.9) | 49 (20.4) | 0.127 |
|  | Overweight | 203 (49.9) | 30 (44.1) |  | 122 (33.9) | 89 (37.1) |  |
|  | Obesity | 115 (28.3) | 23 (33.8) |  | 117 (32.5) | 87 (36.2) |  |
|  | Missing | 13 (3.2) | 0 (0.0) |  | 17 (4.7) | 15 (6.2) |  |
| **Smoking status** | Current | 37 (9.1) | 5 (7.4) | 0.069 | 29 (8.1) | 24 (10.0) | 0.712 |
|  | Former | 247 (60.7) | 51 (75.0) |  | 177 (49.2) | 115 (47.9) |  |
|  | Never | 123 (30.2) | 12 (17.6) |  | 154 (42.8) | 101 (42.1) |  |
|  | Missing | 0 (0.0) | 0 (0.0) |  | 0 (0.0) | 0 (0.0) |  |
| **Drinking patterns** | Heavy | 133 (32.7) | 19 (27.9) | 0.304 | 63 (17.5) | 24 (10.0) | <0.001 |
|  | Moderate | 175 (43.0) | 24 (35.3) |  | 158 (43.9) | 51 (21.2) |  |
|  | Light | 55 (13.5) | 13 (19.1) |  | 87 (24.2) | 83 (34.6) |  |
|  | Never | 36 (8.8) | 10 (14.7) |  | 48 (13.3) | 67 (27.9) |  |
|  | Missing | 8 (2.0) | 2 (2.9) |  | 4 (1.1) | 15 (6.2) |  |
| **Internet use** | No | 139 (34.2) | 34 (50.0) | 0.013 | 152 (42.2) | 154 (64.2) | <0.001 |
|  | Yes | 260 (63.9) | 31 (45.6) |  | 205 (56.9) | 71 (29.6) |  |
|  | Missing | 8 (2.0) | 3 (4.4) |  | 3 (0.8) | 15 (6.2) |  |
| **Comorbidities** | Mild | 309 (75.9) | 51 (75.0) | 0.911 | 288 (80.0) | 168 (70.0) | 0.008 |
|  | Moderate | 79 (19.4) | 13 (19.1) |  | 63 (17.5) | 57 (23.8) |  |
|  | Severe | 19 (4.7) | 4 (5.9) |  | 9 (2.5) | 15 (6.2) |  |
|  | Missing | 0 (0.0) | 0 (0.0) |  | 0 (0.0) | 0 (0.0) |  |
| **Baseline cognition** | ≤25% | 117 (28.7) | 34 (50.0) | 0.001 | 85 (23.6) | 103 (42.9) | <0.001 |
|  | 25~50% | 104 (25.6) | 19 (27.9) |  | 77 (21.4) | 53 (22.1) |  |
|  | 50%~75% | 103 (25.3) | 7 (10.3) |  | 78 (21.7) | 47 (19.6) |  |
|  | ＞75% | 83 (20.4) | 8 (11.8) |  | 120 (33.3) | 37 (15.4) |  |
|  | Missing | 0 (0.0) | 0 (0.0) |  | 0 (0.0) | 0 (0.0) |  |

*Note: Data are N (%) for categorical variables or mean (SD) for continuous variables.*

*ELSA, the English Longitudinal Study of Ageing; BMI, body mass index.*

***Table S6: Characteristics of MHAS participants by sex***

|  | | **Male (*n*=656)** | | ***p* value** | **Female (*n*=897)** | | ***p* value** |
| --- | --- | --- | --- | --- | --- | --- | --- |
|  |  | **Married**  ***n*=568 (86.6)** | **Widowed**  ***n*=88 (13.4)** |  | **Married**  ***n*=578 (64.4)** | **Widowed**  ***n*=319 (35.6)** |  |
| **Age (in years)** |  | 65.06 (7.83) | 73.23 (8.74) | <0.001 | 62.41 (6.87) | 69.01 (8.97) | <0.001 |
| **Minority status** | Urban community | 382 (67.3) | 58 (65.9) | 0.898 | 393 (68.0) | 251 (78.7) | 0.001 |
|  | Rural village | 186 (32.7) | 30 (34.1) |  | 185 (32.0) | 68 (21.3) |  |
|  | Missing | 0 (0.0) | 0 (0.0) |  | 0 (0.0) | 0 (0.0) |  |
| **Educational attainment** | Less than upper secondary | 463 (81.5) | 78 (88.6) | 0.265 | 530 (91.7) | 296 (92.8) | 0.843 |
|  | Upper secondary or vocational training | 20 (3.5) | 3 (3.4) |  | 11 (1.9) | 5 (1.6) |  |
|  | Tertiary | 71 (12.5) | 7 (8.0) |  | 36 (6.2) | 18 (5.6) |  |
|  | Missing | 14 (2.5) | 0 (0.0) |  | 1 (0.2) | 0 (0.0) |  |
| **Household wealth level** | Low | 201 (35.4) | 50 (56.8) | <0.001 | 204 (35.3) | 159 (49.8) | <0.001 |
|  | Normal | 215 (37.9) | 28 (31.8) |  | 215 (37.2) | 111 (34.8) |  |
|  | High | 152 (26.8) | 10 (11.4) |  | 159 (27.5) | 49 (15.4) |  |
|  | Missing | 0 (0.0) | 0 (0.0) |  | 0 (0.0) | 0 (0.0) |  |
| **BMI** | Underweight/Normal | 186 (32.7) | 38 (43.2) | 0.009 | 145 (25.1) | 100 (31.3) | 0.042 |
|  | Overweight | 247 (43.5) | 42 (47.7) |  | 235 (40.7) | 115 (36.1) |  |
|  | Obesity | 125 (22.0) | 6 (6.8) |  | 175 (30.3) | 83 (26.0) |  |
|  | Missing | 10 (1.8) | 2 (2.3) |  | 23 (4.0) | 21 (6.6) |  |
| **Smoking status** | Current | 101 (17.8) | 14 (15.9) | 0.310 | 29 (5.0) | 25 (7.8) | 0.166 |
|  | Former | 257 (45.2) | 34 (38.6) |  | 76 (13.1) | 47 (14.7) |  |
|  | Never | 210 (37.0) | 40 (45.5) |  | 473 (81.8) | 247 (77.4) |  |
|  | Missing | 0 (0.0) | 0 (0.0) |  | 0 (0.0) | 0 (0.0) |  |
| **Drinking patterns** | Heavy | 20 (3.5) | 5 (5.7) | 0.241 | 1 (0.2) | 1 (0.3) | 0.472 |
|  | Moderate | 136 (23.9) | 16 (18.2) |  | 41 (7.1) | 16 (5.0) |  |
|  | Light | 64 (11.3) | 15 (17.0) |  | 32 (5.5) | 23 (7.2) |  |
|  | Never | 348 (61.3) | 52 (59.1) |  | 504 (87.2) | 279 (87.5) |  |
|  | Missing | 0 (0.0) | 0 (0.0) |  | 0 (0.0) | 0 (0.0) |  |
| **Internet use** | No | 260 (45.8) | 45 (51.1) | 0.410 | 217 (37.5) | 138 (43.3) | 0.109 |
|  | Yes | 308 (54.2) | 43 (48.9) |  | 361 (62.5) | 181 (56.7) |  |
|  | Missing | 0 (0.0) | 0 (0.0) |  | 0 (0.0) | 0 (0.0) |  |
| **Comorbidities** | Mild | 443 (78.0) | 67 (76.1) | 0.463 | 434 (75.1) | 228 (71.5) | 0.142 |
|  | Moderate | 100 (17.6) | 19 (21.6) |  | 121 (20.9) | 69 (21.6) |  |
|  | Severe | 25 (4.4) | 2 (2.3) |  | 23 (4.0) | 22 (6.9) |  |
|  | Missing | 0 (0.0) | 0 (0.0) |  | 0 (0.0) | 0 (0.0) |  |
| **Baseline cognition** | ≤25% | 157 (27.6) | 39 (44.3) | 0.013 | 119 (20.6) | 81 (25.4) | 0.116 |
|  | 25~50% | 157 (27.6) | 21 (23.9) |  | 128 (22.1) | 75 (23.5) |  |
|  | 50%~75% | 139 (24.5) | 17 (19.3) |  | 163 (28.2) | 92 (28.8) |  |
|  | ＞75% | 115 (20.2) | 11 (12.5) |  | 168 (29.1) | 71 (22.3) |  |
|  | Missing | 0 (0.0) | 0 (0.0) |  | 0 (0.0) | 0 (0.0) |  |

*Note: Data are N (%) for categorical variables or mean (SD) for continuous variables.*

*MHAS, the Mexican Health and Aging Study; BMI, body mass index.*

***Table S7: Characteristics of CHARLS participants by sex***

|  | | **Male (*n*=4446)** | | ***p* value** | **Female (*n*=4311)** | | ***p* value** |
| --- | --- | --- | --- | --- | --- | --- | --- |
|  |  | **Married**  ***n*=4071(91.6)** | **Widowed**  ***n*=375(8.4)** |  | **Married**  ***n*=3455(80.1)** | **Widowed**  ***n*=856(19.9)** |  |
| **Age (in years)** |  | 61.20 (6.29) | 65.43 (7.47) | <0.001 | 60.18 (5.56) | 65.43 (7.30) | <0.001 |
| **Minority status** | Urban community | 1611 (39.6) | 112 (29.9) | <0.001 | 1413 (40.9) | 367 (42.9) | 0.311 |
|  | Rural village | 2460 (60.4) | 263 (70.1) |  | 2042 (59.1) | 489 (57.1) |  |
|  | Missing | 0 (0.0) | 0 (0.0) |  | 0 (0.0) | 0 (0.0) |  |
| **Educational attainment** | Less than upper secondary | 3465 (85.1) | 341 (90.9) | 0.005 | 3215 (93.1) | 818 (95.6) | 0.028 |
|  | Upper secondary or vocational training | 512 (12.6) | 26 (6.9) |  | 211 (6.1) | 33 (3.9) |  |
|  | Tertiary | 94 (2.3) | 8 (2.1) |  | 29 (0.8) | 5 (0.6) |  |
|  | Missing | 0 (0.0) | 0 (0.0) |  | 0 (0.0) | 0 (0.0) |  |
| **Household wealth level** | Low | 1356 (33.3) | 171 (45.6) | <0.001 | 1212 (35.1) | 427 (49.9) | <0.001 |
|  | Normal | 1228 (30.2) | 98 (26.1) |  | 1035 (30.0) | 206 (24.1) |  |
|  | High | 1404 (34.5) | 99 (26.4) |  | 1155 (33.4) | 187 (21.8) |  |
|  | Missing | 83 (2.0) | 7 (1.9) |  | 53 (1.5) | 36 (4.2) |  |
| **BMI** | Underweight/Normal | 2769 (68.0) | 290 (77.3) | 0.002 | 1997 (57.8) | 549 (64.1) | 0.009 |
|  | Overweight | 958 (23.5) | 63 (16.8) |  | 1076 (31.1) | 231 (27.0) |  |
|  | Obesity | 89 (2.2) | 4 (1.1) |  | 225 (6.5) | 44 (5.1) |  |
|  | Missing | 255 (6.3) | 18 (4.8) |  | 157 (4.5) | 32 (3.7) |  |
| **Smoking status** | Current | 2290 (56.3) | 224 (59.7) | 0.328 | 209 (6.0) | 73 (8.5) | <0.001 |
|  | Former | 714 (17.5) | 63 (16.8) |  | 68 (2.0) | 25 (2.9) |  |
|  | Never | 1046 (25.7) | 88 (23.5) |  | 3177 (92.0) | 755 (88.2) |  |
|  | Missing | 21 (0.5) | 0 (0.0) |  | 1 (0.0) | 3 (0.4) |  |
| **Drinking patterns** | Heavy | 1171 (28.8) | 103 (27.5) | 0.466 | 99 (2.9) | 29 (3.4) | 0.007 |
|  | Moderate | 312 (7.7) | 23 (6.1) |  | 49 (1.4) | 16 (1.9) |  |
|  | Light | 1211 (29.7) | 113 (30.1) |  | 377 (10.9) | 101 (11.8) |  |
|  | Never | 1361 (33.4) | 136 (36.3) |  | 2928 (84.7) | 705 (82.4) |  |
|  | Missing | 16 (0.4) | 0 (0.0) |  | 2 (0.1) | 5 (0.6) |  |
| **Internet use** | No | 3166 (77.8) | 309 (82.4) | 0.025 | 2829 (81.9) | 720 (84.1) | 0.277 |
|  | Yes | 114 (2.8) | 3 (0.8) |  | 64 (1.9) | 12 (1.4) |  |
|  | Missing | 791 (19.4) | 63 (16.8) |  | 562 (16.3) | 124 (14.5) |  |
| **Comorbidities** | Mild | 3482 (85.5) | 328 (87.5) | 0.706 | 2884 (83.5) | 701 (81.9) | 0.450 |
|  | Moderate | 302 (7.4) | 24 (6.4) |  | 328 (9.5) | 97 (11.3) |  |
|  | Severe | 59 (1.4) | 6 (1.6) |  | 69 (2.0) | 17 (2.0) |  |
|  | Missing | 228 (5.6) | 17 (4.5) |  | 174 (5.0) | 41 (4.8) |  |
| **Baseline cognition** | ≤25% | 1223 (30.0) | 135 (36.0) | 0.007 | 1209 (35.0) | 339 (39.6) | 0.006 |
|  | 25~50% | 944 (23.2) | 100 (26.7) |  | 797 (23.1) | 218 (25.5) |  |
|  | 50%~75% | 929 (22.8) | 70 (18.7) |  | 705 (20.4) | 149 (17.4) |  |
|  | ＞75% | 905 (22.2) | 68 (18.1) |  | 715 (20.7) | 143 (16.7) |  |
|  | Missing | 70 (1.7) | 2 (0.5) |  | 29 (0.8) | 7 (0.8) |  |

*Note: Data are N (%) for categorical variables or mean (SD) for continuous variables.*

*CHARLS, the China Health Retirement Longitudinal Study; BMI, body mass index*

***Table S8: Characteristics of LASI participants by sex***

|  | | **Male (*n*=1794)** | | ***p* value** | **Female (*n*=2130)** | | ***p* value** |
| --- | --- | --- | --- | --- | --- | --- | --- |
|  |  | **Married**  ***n*=1534(85.5)** | **Widowed**  ***n*=260(14.5)** |  | **Married**  ***n*=1077(50.6)** | **Widowed**  ***n*=1053(49.4)** |  |
| **Age (in years)** |  | 68.73 (6.73) | 73.08 (8.83) | <0.001 | 65.50 (5.25) | 71.44 (8.37) | <0.001 |
| **Minority status** | Urban community | 533 (34.7) | 90 (34.6) | 1.000 | 394 (36.6) | 454 (43.1) | 0.002 |
|  | Rural village | 1001 (65.3) | 170 (65.4) |  | 683 (63.4) | 599 (56.9) |  |
|  | Missing | 0 (0.0) | 0 (0.0) |  | 0 (0.0) | 0 (0.0) |  |
| **Educational attainment** | Less than upper secondary | 948 (61.8) | 178 (68.5) | 0.057 | 890 (82.6) | 951 (90.3) | <0.001 |
|  | Upper secondary or vocational training | 472 (30.8) | 71 (27.3) |  | 163 (15.1) | 94 (8.9) |  |
|  | Tertiary | 114 (7.4) | 11 (4.2) |  | 24 (2.2) | 8 (0.8) |  |
|  | Missing | 0 (0.0) | 0 (0.0) |  | 0 (0.0) | 0 (0.0) |  |
| **Household wealth level** | Low | 449 (29.3) | 80 (30.8) | 0.432 | 322 (29.9) | 380 (36.1) | <0.001 |
|  | Normal | 489 (31.9) | 87 (33.5) |  | 320 (29.7) | 337 (32.0) |  |
|  | High | 498 (32.5) | 83 (31.9) |  | 369 (34.3) | 279 (26.5) |  |
|  | Missing | 98 (6.4) | 10 (3.8) |  | 66 (6.1) | 57 (5.4) |  |
| **BMI** | Underweight/Normal | 1094 (71.3) | 186 (71.5) | 0.543 | 620 (57.6) | 663 (63.0) | <0.001 |
|  | Overweight | 270 (17.6) | 52 (20.0) |  | 254 (23.6) | 207 (19.7) |  |
|  | Obesity | 38 (2.5) | 5 (1.9) |  | 136 (12.6) | 76 (7.2) |  |
|  | Missing | 132 (8.6) | 17 (6.5) |  | 67 (6.2) | 107 (10.2) |  |
| **Smoking status** | Current | 727 (47.4) | 118 (45.4) | 0.919 | 219 (20.3) | 253 (24.0) | 0.227 |
|  | Former | 183 (11.9) | 31 (11.9) |  | 24 (2.2) | 21 (2.0) |  |
|  | Never | 610 (39.8) | 109 (41.9) |  | 830 (77.1) | 776 (73.7) |  |
|  | Missing | 14 (0.9) | 2 (0.8) |  | 4 (0.4) | 3 (0.3) |  |
| **Drinking patterns** | Heavy | 40 (2.6) | 7 (2.7) | 0.968 | 4 (0.4) | 3 (0.3) | 0.188 |
|  | Moderate | 45 (2.9) | 8 (3.1) |  | 2 (0.2) | 3 (0.3) |  |
|  | Light | 374 (24.4) | 58 (22.3) |  | 11 (1.0) | 24 (2.3) |  |
|  | Never | 1062 (69.2) | 185 (71.2) |  | 1056 (98.1) | 1021 (97.0) |  |
|  | Missing | 13 (0.8) | 2 (0.8) |  | 4 (0.4) | 2 (0.2) |  |
| **Internet use** | No | 1448 (94.4) | 253 (97.3) | 0.128 | 1043 (96.8) | 1032 (98.0) | 0.237 |
|  | Yes | 60 (3.9) | 4 (1.5) |  | 20 (1.9) | 12 (1.1) |  |
|  | Missing | 26 (1.7) | 3 (1.2) |  | 14 (1.3) | 9 (0.9) |  |
| **Comorbidities** | Mild | 1283 (83.6) | 225 (86.5) | 0.504 | 908 (84.3) | 897 (85.2) | 0.955 |
|  | Moderate | 216 (14.1) | 32 (12.3) |  | 150 (13.9) | 138 (13.1) |  |
|  | Severe | 29 (1.9) | 2 (0.8) |  | 18 (1.7) | 17 (1.6) |  |
|  | Missing | 6 (0.4) | 1 (0.4) |  | 1 (0.1) | 1 (0.1) |  |
| **Baseline cognition** | ≤25% | 364 (23.7) | 80 (30.8) | 0.004 | 237 (22.0) | 401 (38.1) | <0.001 |
|  | 25~50% | 453 (29.5) | 90 (34.6) |  | 377 (35.0) | 316 (30.0) |  |
|  | 50%~75% | 312 (20.3) | 44 (16.9) |  | 212 (19.7) | 174 (16.5) |  |
|  | ＞75% | 392 (25.6) | 43 (16.5) |  | 246 (22.8) | 139 (13.2) |  |
|  | Missing | 13 (0.8) | 3 (1.2) |  | 5 (0.5) | 23 (2.2) |  |

*Note: Data are N (%) for categorical variables or mean (SD) for continuous variables.*

*LASI, the Longitudinal Aging Study in India; BMI, body mass index.*

***Table S9: Characteristics of HAALSI participants by sex***

|  | | **Male (*n*=201)** | | ***p* value** | **Female (*n*=369)** | | ***p* value** |
| --- | --- | --- | --- | --- | --- | --- | --- |
|  |  | **Married**  ***n*=161 (80.1)** | **Widowed**  ***n*=40 (19.9)** |  | **Married**  ***n*=102 (27.6)** | **Widowed**  ***n*=267 (72.4)** |  |
| **Age (in years)** |  | 64.06 (10.15) | 67.75 (11.82) | 0.048 | 57.52 (8.56) | 67.99 (11.41) | <0.001 |
| **Educational attainment** | Less than upper secondary | 151 (93.8) | 39 (97.5) | 0.587 | 84 (82.4) | 250 (93.6) | 0.003 |
|  | Upper secondary or vocational training | 7 (4.3) | 1 (2.5) |  | 14 (13.7) | 15 (5.6) |  |
|  | Tertiary | 3 (1.9) | 0 (0.0) |  | 4 (3.9) | 2 (0.7) |  |
|  | Missing | 0 (0.0) | 0 (0.0) |  | 0 (0.0) | 0 (0.0) |  |
| **Household wealth level** | Low | 45 (28.0) | 18 (45.0) | 0.062 | 15 (14.7) | 91 (34.1) | <0.001 |
|  | Normal | 44 (27.3) | 13 (32.5) |  | 29 (28.4) | 81 (30.3) |  |
|  | High | 46 (28.6) | 5 (12.5) |  | 39 (38.2) | 77 (28.8) |  |
|  | Missing | 26 (16.1) | 4 (10.0) |  | 19 (18.6) | 18 (6.7) |  |
| **BMI** | Underweight/Normal | 68 (42.2) | 28 (70.0) | 0.002 | 21 (20.6) | 89 (33.3) | 0.118 |
|  | Overweight | 53 (32.9) | 12 (30.0) |  | 34 (33.3) | 77 (28.8) |  |
|  | Obesity | 30 (18.6) | 0 (0.0) |  | 40 (39.2) | 84 (31.5) |  |
|  | Missing | 10 (6.2) | 0 (0.0) |  | 7 (6.9) | 17 (6.4) |  |
| **Smoking status** | Current | 25 (15.5) | 6 (15.0) | 0.711 | 1 (1.0) | 0 (0.0) | 0.255 |
|  | Former | 46 (28.6) | 9 (22.5) |  | 2 (2.0) | 4 (1.5) |  |
|  | Never | 90 (55.9) | 25 (62.5) |  | 99 (97.1) | 263 (98.5) |  |
|  | Missing | 0 (0.0) | 0 (0.0) |  | 0 (0.0) | 0 (0.0) |  |
| **Drinking patterns** | Heavy | 14 (8.7) | 5 (12.5) | 0.738 | 2 (2.0) | 6 (2.2) | 0.240 |
|  | Moderate | 18 (11.2) | 3 (7.5) |  | 2 (2.0) | 7 (2.6) |  |
|  | Light | 80 (49.7) | 18 (45.0) |  | 21 (20.6) | 82 (30.7) |  |
|  | Never | 49 (30.4) | 14 (35.0) |  | 77 (75.5) | 172 (64.4) |  |
|  | Missing | 0 (0.0) | 0 (0.0) |  | 0 (0.0) | 0 (0.0) |  |
| **Internet use** | No | 17 (10.6) | 10 (25.0) | 0.056 | 11 (10.8) | 36 (13.5) | 0.431 |
|  | Yes | 130 (80.7) | 27 (67.5) |  | 87 (85.3) | 213 (79.8) |  |
|  | Missing | 14 (8.7) | 3 (7.5) |  | 4 (3.9) | 18 (6.7) |  |
| **Comorbidities** | Mild | 143 (88.8) | 36 (90.0) | 0.278 | 81 (79.4) | 212 (79.4) | 0.799 |
|  | Moderate | 8 (5.0) | 4 (10.0) |  | 11 (10.8) | 34 (12.7) |  |
|  | Severe | 2 (1.2) | 0 (0.0) |  | 1 (1.0) | 4 (1.5) |  |
|  | Missing | 8 (5.0) | 0 (0.0) |  | 9 (8.8) | 17 (6.4) |  |
| **Baseline cognition** | ≤25% | 56 (34.8) | 19 (47.5) | 0.305 | 25 (24.5) | 118 (44.2) | 0.006 |
|  | 25~50% | 19 (11.8) | 7 (17.5) |  | 16 (15.7) | 33 (12.4) |  |
|  | 50%~75% | 53 (32.9) | 10 (25.0) |  | 41 (40.2) | 76 (28.5) |  |
|  | ＞75% | 32 (19.9) | 4 (10.0) |  | 20 (19.6) | 36 (13.5) |  |
|  | Missing | 1 (0.6) | 0 (0.0) |  | 0 (0.0) | 4 (1.5) |  |

*Note: Data are N (%) for categorical variables or mean (SD) for continuous variables.*

*For HAALSI, the question on minority status was unavailable.*

*HAALSI, the Health and Aging in Africa: A Longitudinal Study of an INDEPTH Community in South Africa; BMI, body mass index.*

**Ⅵ. Meta-analysis of outcomes**

***Table S10: Overall meta-analysis estimates, 95% confidence interval, and I^2^ heterogeneity of specific-domain***

|  | ***β* (95% CI)** | ***p*-value** | ***I*^2^** |
| --- | --- | --- | --- |
| **Orientation** | 0.00 (-0.06, 0.07) | 0.8785 | 95.21% |
| **Memory** | -0.03(-0.08, 0.02) | 0.2771 | 93.97% |
| **Executive function** | -0.14(-0.19, -0.08) | <.0001 | 94.31% |
| **Language** | -0.08(-0.19, 0.04) | 0.1937 | 98.41% |

***Table S11: Male meta-analysis estimates, 95% confidence interval, and I^2^ heterogeneity of specific-domain***

|  | ***β* (95% CI)** | ***p*-value** | ***I*^2^** |
| --- | --- | --- | --- |
| **Orientation** | 0.00 (-0.06, 0.07) | 0.8785 | 95.21% |
| **Memory** | -0.03(-0.08, 0.02) | 0.2771 | 93.97% |
| **Executive function** | -0.14(-0.19, -0.08) | <.0001 | 94.31% |
| **Language** | -0.08(-0.19, 0.04) | 0.1937 | 98.41% |

***Table S12: Female meta-analysis estimates, 95% confidence interval, and I^2^ heterogeneity of specific-domain***

|  | ***β* (95% CI)** | ***p*-value** | ***I*^2^** |
| --- | --- | --- | --- |
| **Orientation** | -0.03(-0.09, 0.03) | 0.3292 | 97.15% |
| **Memory** | -0.07(-0.14, -0.01) | 0.0282 | 97.65% |
| **Executive function** | -0.05(-0.12, 0.01) | 0.0970 | 98.12% |
| **Language** | -0.01(-0.08, 0.07) | 0.8755 | 98.38% |

*95% CI: 95% confidence interval.*

**Ⅶ. Restricted cubic spline (RCS) regressions**

***Figure S7: Association between age and cognitive function by widowhood status in HRS, MHAS, and CHARLS***

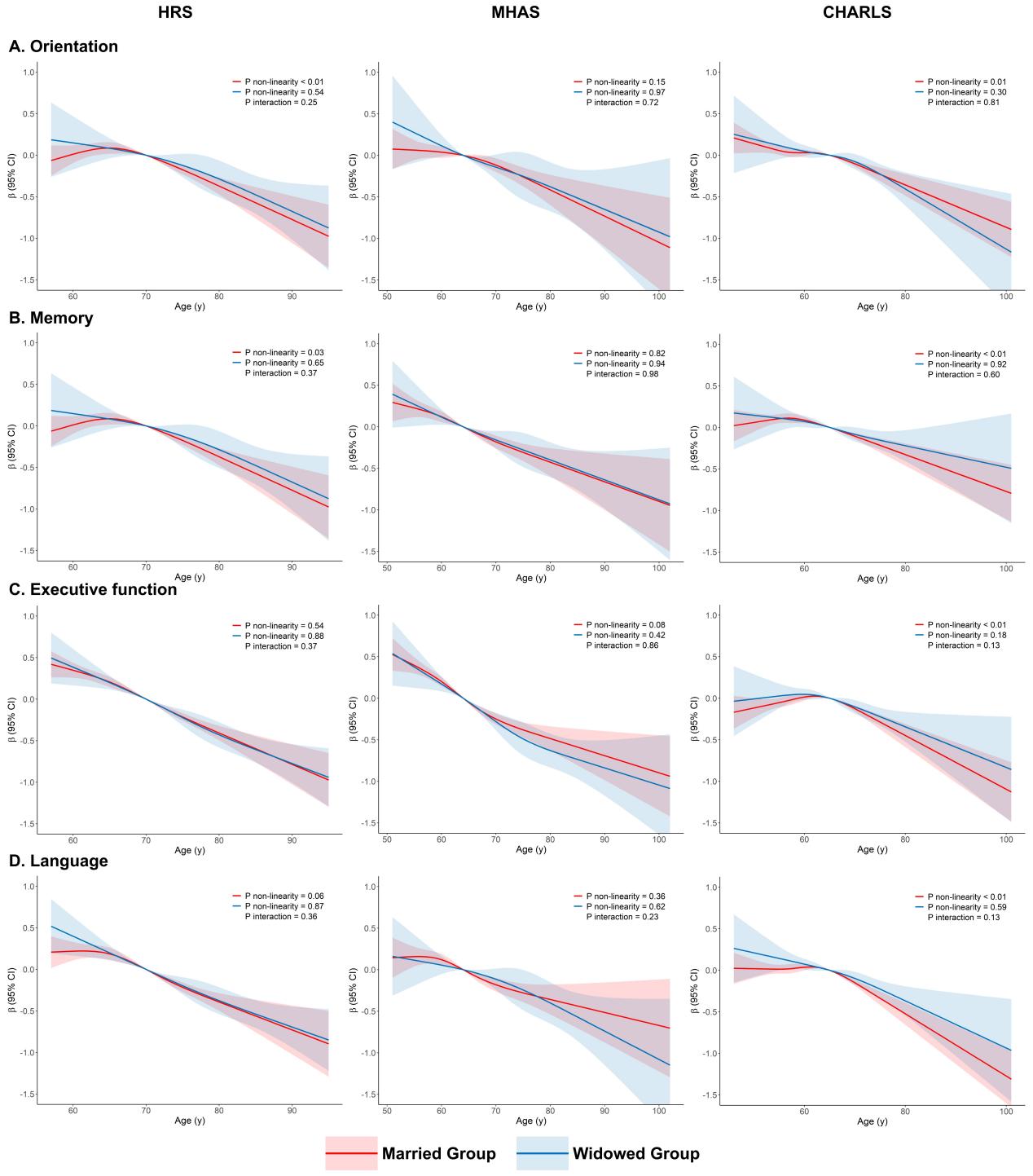

*NOTE: Models adjusted for age, sex, minority status, educational attainment, household wealth level, BMI, smoking status, drinking pattern, internet use, and comorbidities.*

*HRS, the Health and Retirement Study; MHAS, the Mexican Health and Aging Study; CHARLS, the China Health Retirement Longitudinal Study.*

***Figure S8: Association between age and cognitive function by widowhood status in ELSA, LASI, and HAALSI***

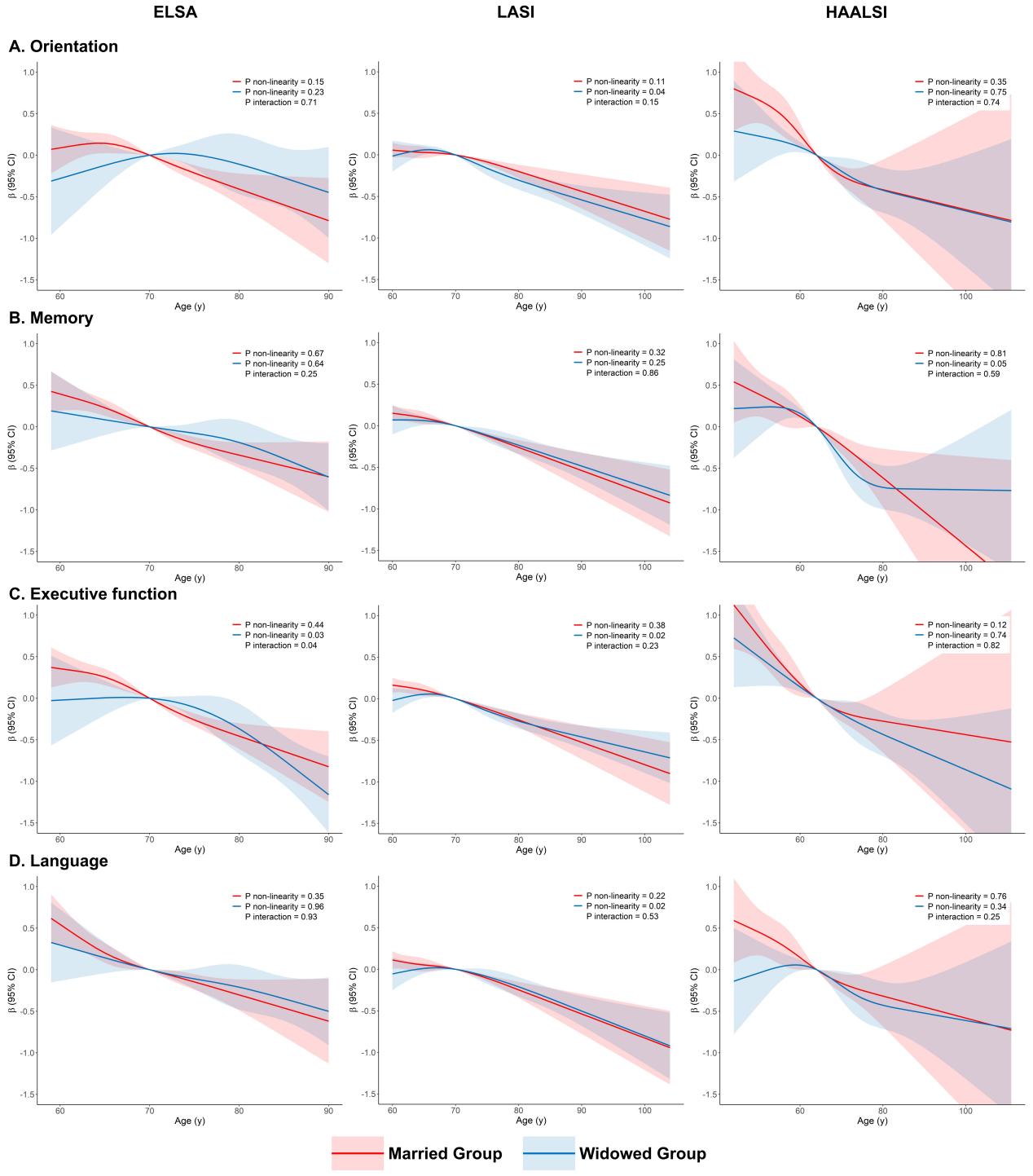

*NOTE: Models adjusted for age, sex, minority status (except for the HAALSI), educational attainment, household wealth level, BMI, smoking status, drinking pattern, internet use, and comorbidities.*

*ELSA, the English Longitudinal Study of Ageing; LASI, the Longitudinal Aging Study in India; HAALSI, the Health and Aging in Africa.*

**Ⅷ. Cognitive correlates in widowed participants**

***Table S13: Individual-characteristics with cognitive function in the HRS widowed participants***

|  | **Events/Observations** | **Orientation** | **Memory** | **Executive function** | **Language** |
| --- | --- | --- | --- | --- | --- |
| **Educational attainment** | | | | | |
| Less than upper secondary | 203/851 | 0 [Reference] | 0 [Reference] | 0 [Reference] | 0 [Reference] |
| Upper secondary or vocational training | 515/851 | 0.17(0, 0.34) | 0.24(0.10, 0.38) | 0.48(0.34, 0.61) | 0.09(-0.06, 0.25) |
| Tertiary | 133/851 | 0.04(-0.2, 0.28) | 0.27(0.07, 0.48) | 0.56(0.36, 0.75) | 0.18(-0.05, 0.40) |
| **Household wealth level** | | | | | |
| Low | 378/851 | 0 [Reference] | 0 [Reference] | 0 [Reference] | 0 [Reference] |
| Normal | 271/851 | 0.15(-0.01, 0.31) | 0.08(-0.06, 0.21) | 0.26(0.14, 0.39) | 0.15(0.01, 0.30) |
| High | 202/851 | 0.12(-0.07, 0.31) | 0.07(-0.09, 0.23) | 0.21(0.06, 0.36) | 0.24(0.07, 0.41) |
| **BMI** | | | | | |
| Underweight/Normal | 200/851 | 0 [Reference] | 0 [Reference] | 0 [Reference] | 0 [Reference] |
| Overweight | 277/851 | 0.17(-0.01, 0.35) | 0.03(-0.12, 0.18) | -0.07(-0.21, 0.07) | -0.03(-0.20, 0.13) |
| Obesity | 374/851 | 0.27(0.10, 0.44) | 0.11(-0.04, 0.25) | 0.03(-0.11, 0.17) | 0.04(-0.12, 0.20) |
| **Smoking status** | | | | | |
| Current | 75/851 | 0 [Reference] | 0 [Reference] | 0 [Reference] | 0 [Reference] |
| Former | 345/851 | 0.04(-0.20, 0.29) | 0.04(-0.17, 0.25) | 0.06(-0.14, 0.26) | 0.04(-0.19, 0.27) |
| Never | 431/851 | 0.11(-0.14, 0.36) | 0.06(-0.15, 0.27) | 0.12(-0.08, 0.32) | 0.12(-0.10, 0.35) |
| **Drinking patterns** | | | | | |
| Heavy | 69/851 | 0 [Reference] | 0 [Reference] | 0 [Reference] | 0 [Reference] |
| Moderate | 172/851 | 0.07(-0.19, 0.34) | 0.15(-0.08, 0.37) | -0.05(-0.27, 0.16) | 0.02(-0.23, 0.26) |
| Light | 139/851 | 0(-0.28, 0.28) | 0.03(-0.20, 0.27) | -0.07(-0.30, 0.16) | -0.12(-0.37, 0.14) |
| Never | 471/851 | -0.05(-0.30, 0.21) | -0.06(-0.28, 0.16) | -0.12(-0.33, 0.09) | -0.25(-0.49, -0.01) |
| **Internet use** | | | | | |
| No | 553/851 | 0 [Reference] | 0 [Reference] | 0 [Reference] | 0 [Reference] |
| Yes | 298/851 | 0.03(-0.13, 0.19) | 0.17(0.04, 0.31) | 0.25(0.12, 0.38) | 0.16(0.02, 0.31) |
| **Comorbidities** | | | | | |
| Mild | 526/851 | 0 [Reference] | 0 [Reference] | 0 [Reference] | 0 [Reference] |
| Moderate | 238/851 | 0.10(-0.05, 0.25) | 0.03(-0.10, 0.15) | 0.02(-0.11, 0.14) | 0(-0.13, 0.14) |
| Severe | 87/851 | -0.10(-0.32, 0.12) | 0.08(-0.10, 0.27) | -0.11(-0.29, 0.07) | -0.05(-0.25, 0.15) |

*HRS, the Health and Retirement Study; BMI, body mass index.*

***Table S14: Individual-characteristics with cognitive function in the ELSA widowed participants***

|  | **Events/Observations** | **Orientation** | **Memory** | **Executive function** | **Language** |
| --- | --- | --- | --- | --- | --- |
| **Educational attainment** | | | | | |
| Less than upper secondary | 177/308 | 0 [Reference] | 0 [Reference] | 0 [Reference] | 0 [Reference] |
| Upper secondary or vocational training | 113/308 | 0.26(0.02, 0.50) | 0.12(-0.08, 0.31) | 0.22(0.02, 0.43) | 0.08(-0.14, 0.29) |
| Tertiary | 18/308 | 0.26(-0.24, 0.77) | 0.10(-0.30, 0.51) | 0.28(-0.15, 0.72) | 0.27(-0.18, 0.72) |
| **Household wealth level** | | | | | |
| Low | 196/308 | 0 [Reference] | 0 [Reference] | 0 [Reference] | 0 [Reference] |
| Normal | 74/308 | 0.23(-0.03, 0.50) | 0.15(-0.06, 0.36) | 0.16(-0.07, 0.38) | -0.03(-0.26, 0.21) |
| High | 38/308 | 0(-0.35, 0.35) | 0.14(-0.14, 0.42) | -0.06(-0.36, 0.24) | 0.02(-0.29, 0.33) |
| **BMI** | | | | | |
| Underweight/Normal | 67/308 | 0 [Reference] | 0 [Reference] | 0 [Reference] | 0 [Reference] |
| Overweight | 127/308 | -0.04(-0.32, 0.25) | 0.21(-0.02, 0.44) | 0.10(-0.14, 0.35) | 0.28(0.03, 0.54) |
| Obesity | 114/308 | 0.02(-0.28, 0.32) | 0.23(-0.01, 0.47) | 0.08(-0.18, 0.33) | 0.20(-0.07, 0.46) |
| **Smoking status** | | | | | |
| Current | 29/308 | 0 [Reference] | 0 [Reference] | 0 [Reference] | 0 [Reference] |
| Former | 166/308 | -0.20(-0.59, 0.19) | -0.28(-0.60, 0.03) | 0.11(-0.23, 0.45) | -0.13(-0.48, 0.22) |
| Never | 113/308 | -0.03(-0.43, 0.37) | -0.32(-0.64, 0) | 0.18(-0.16, 0.53) | 0(-0.35, 0.35) |
| **Drinking patterns** | | | | | |
| Heavy | 46/308 | 0 [Reference] | 0 [Reference] | 0 [Reference] | 0 [Reference] |
| Moderate | 75/308 | 0.25(-0.11, 0.61) | 0.11(-0.19, 0.40) | 0.04(-0.28, 0.35) | 0.35(0.02, 0.67) |
| Light | 103/308 | 0.14(-0.22, 0.49) | 0.04(-0.25, 0.33) | -0.04(-0.35, 0.27) | 0.17(-0.15, 0.48) |
| Never | 84/308 | 0.26(-0.12, 0.64) | 0.07(-0.23, 0.38) | -0.07(-0.40, 0.25) | 0.06(-0.27, 0.40) |
| **Internet use** | | | | | |
| No | 204/308 | 0 [Reference] | 0 [Reference] | 0 [Reference] | 0 [Reference] |
| Yes | 104/308 | 0.07(-0.21, 0.36) | 0.27(0.05, 0.50) | 0.41(0.17, 0.66) | 0.24(-0.01, 0.49) |
| **Comorbidities** | | | | | |
| Mild | 219/308 | 0 [Reference] | 0 [Reference] | 0 [Reference] | 0 [Reference] |
| Moderate | 70/308 | -0.14(-0.41, 0.12) | -0.04(-0.26, 0.17) | -0.15(-0.37, 0.08) | -0.10(-0.34, 0.13) |
| Severe | 19/308 | -0.27(-0.73, 0.19) | -0.07(-0.44, 0.30) | -0.08(-0.47, 0.32) | -0.20(-0.61, 0.21) |

*ELSA, the English Longitudinal Study of Ageing; BMI, body mass index.*

***Table S15: Individual-characteristics with cognitive function in the MHAS widowed participants***

|  | **Events/Observations** | **Orientation** | **Memory** | **Executive function** | **Language** |
| --- | --- | --- | --- | --- | --- |
| **Educational attainment** | | | | | |
| Less than upper secondary | 374/407 | 0 [Reference] | 0 [Reference] | 0 [Reference] | 0 [Reference] |
| Upper secondary or vocational training | 8/407 | 0.07(-0.59, 0.73) | 0.37(-0.19, 0.94) | 0.54(0, 1.08) | 0.56(-0.07, 1.19) |
| Tertiary | 25/407 | 0.54(0.15, 0.93) | 0.79(0.45, 1.13) | 1.14(0.82, 1.47) | 0.51(0.13, 0.89) |
| **Household wealth level** | | | | | |
| Low | 209/407 | 0 [Reference] | 0 [Reference] | 0 [Reference] | 0 [Reference] |
| Normal | 139/407 | 0.05(-0.14, 0.25) | -0.05(-0.22, 0.12) | 0.19(0.02, 0.35) | 0.11(-0.08, 0.30) |
| High | 59/407 | 0.08(-0.21, 0.36) | -0.03(-0.27, 0.22) | 0.14(-0.09, 0.37) | 0.36(0.09, 0.64) |
| **BMI** | | | | | |
| Underweight/Normal | 148/407 | 0 [Reference] | 0 [Reference] | 0 [Reference] | 0 [Reference] |
| Overweight | 165/407 | 0.12(-0.09, 0.32) | 0.02(-0.16, 0.19) | 0.17(0, 0.33) | 0.19(-0.01, 0.38) |
| Obesity | 94/407 | 0.22(-0.03, 0.47) | 0.23(0.01, 0.44) | 0.24(0.03, 0.44) | 0.27(0.02, 0.51) |
| **Smoking status** | | | | | |
| Current | 39/407 | 0 [Reference] | 0 [Reference] | 0 [Reference] | 0 [Reference] |
| Former | 81/407 | 0.10(-0.26, 0.45) | 0.04(-0.27, 0.34) | -0.03(-0.31, 0.26) | -0.13(-0.47, 0.21) |
| Never | 287/407 | 0.20(-0.12, 0.51) | -0.08(-0.35, 0.19) | -0.14(-0.39, 0.12) | -0.17(-0.47, 0.13) |
| **Drinking patterns** | | | | | |
| Heavy | 6/407 | 0 [Reference] | 0 [Reference] | 0 [Reference] | 0 [Reference] |
| Moderate | 32/407 | -0.10(-0.91, 0.71) | -0.44(-1.13, 0.26) | 0.07(-0.59, 0.73) | -0.20(-0.97, 0.58) |
| Light | 38/407 | -0.13(-0.93, 0.66) | -0.27(-0.96, 0.41) | -0.01(-0.66, 0.64) | -0.18(-0.94, 0.59) |
| Never | 331/407 | -0.20(-0.96, 0.56) | -0.50(-1.15, 0.16) | -0.04(-0.66, 0.58) | -0.24(-0.97, 0.50) |
| **Internet use** | | | | | |
| No | 183/407 | 0 [Reference] | 0 [Reference] | 0 [Reference] | 0 [Reference] |
| Yes | 224/407 | 0.18(-0.01, 0.36) | 0.19(0.03, 0.34) | 0.29(0.14, 0.44) | 0.39(0.22, 0.57) |
| **Comorbidities** | | | | | |
| Mild | 295/407 | 0 [Reference] | 0 [Reference] | 0 [Reference] | 0 [Reference] |
| Moderate | 88/407 | 0.20(-0.03, 0.42) | 0.13(-0.06, 0.32) | -0.03(-0.21, 0.15) | 0.12(-0.10, 0.33) |
| Severe | 24/407 | -0.24(-0.62, 0.14) | 0.25(-0.07, 0.58) | -0.15(-0.46, 0.16) | 0.12(-0.25, 0.48) |

*MHAS, the Mexican Health and Aging Study; BMI, body mass index.*

***Table S16: Individual-characteristics with cognitive function in the CHARLS widowed participants***

|  | **Events/Observations** | **Orientation** | **Memory** | **Executive function** | **Language** |
| --- | --- | --- | --- | --- | --- |
| **Educational attainment** | | | | | |
| Less than upper secondary | 1159/1231 | 0 [Reference] | 0 [Reference] | 0 [Reference] | 0 [Reference] |
| Upper secondary or vocational training | 59/1231 | 0.43(0.19, 0.67) | 0.64(0.39, 0.89) | 0.39(0.13, 0.65) | 0.60(0.35, 0.85) |
| Tertiary | 13/1231 | 0.13(-0.36, 0.63) | 0.32(-0.20, 0.83) | 0.38(-0.15, 0.91) | 0.59(0.08, 1.09) |
| **Household wealth level** | | | | | |
| Low | 613/1231 | 0 [Reference] | 0 [Reference] | 0 [Reference] | 0 [Reference] |
| Normal | 317/1231 | 0.08(-0.04, 0.20) | 0.06(-0.07, 0.18) | 0.01(-0.12, 0.14) | 0.04(-0.08, 0.16) |
| High | 301/1231 | 0.06(-0.07, 0.18) | 0.18(0.05, 0.31) | 0.07(-0.06, 0.21) | 0.14(0.01, 0.26) |
| **BMI** | | | | | |
| Underweight/Normal | 870/1231 | 0 [Reference] | 0 [Reference] | 0 [Reference] | 0 [Reference] |
| Overweight | 310/1231 | 0.04(-0.08, 0.15) | 0.03(-0.10, 0.15) | 0.01(-0.12, 0.13) | 0.14(0.02, 0.26) |
| Obesity | 51/1231 | 0.30(0.05, 0.56) | -0.03(-0.30, 0.23) | 0.22(-0.05, 0.49) | 0.19(-0.07, 0.44) |
| **Smoking status** | | | | | |
| Current | 297/1231 | 0 [Reference] | 0 [Reference] | 0 [Reference] | 0 [Reference] |
| Former | 88/1231 | 0.12(-0.09, 0.34) | -0.02(-0.24, 0.20) | 0.05(-0.17, 0.28) | 0.23(0.02, 0.44) |
| Never | 846/1231 | 0.11(-0.04, 0.26) | -0.03(-0.18, 0.13) | -0.03(-0.19, 0.13) | 0.02(-0.13, 0.17) |
| **Drinking patterns** | | | | | |
| Heavy | 132/1231 | 0 [Reference] | 0 [Reference] | 0 [Reference] | 0 [Reference] |
| Moderate | 39/1231 | 0.11(-0.21, 0.43) | 0.19(-0.14, 0.52) | 0.21(-0.12, 0.55) | -0.05(-0.37, 0.27) |
| Light | 214/1231 | 0.10(-0.10, 0.29) | 0.09(-0.11, 0.29) | -0.05(-0.26, 0.15) | 0.06(-0.14, 0.25) |
| Never | 846/1231 | 0.07(-0.11, 0.24) | 0.09(-0.10, 0.28) | -0.09(-0.29, 0.10) | 0.05(-0.13, 0.24) |
| **Internet use** | | | | | |
| No | 1210/1231 | 0 [Reference] | 0 [Reference] | 0 [Reference] | 0 [Reference] |
| Yes | 21/1231 | 0.17(-0.23, 0.58) | 0.09(-0.34, 0.51) | 0.49(0.05, 0.92) | 0.68(0.27, 1.10) |
| **Comorbidities** | | | | | |
| Mild | 1075/1231 | 0 [Reference] | 0 [Reference] | 0 [Reference] | 0 [Reference] |
| Moderate | 130/1231 | -0.02(-0.19, 0.14) | 0.08(-0.08, 0.25) | -0.01(-0.18, 0.17) | -0.18(-0.34, -0.02) |
| Severe | 26/1231 | 0.20(-0.14, 0.55) | 0.14(-0.22, 0.50) | 0.40(0.03, 0.76) | 0.21(-0.14, 0.56) |

*CHARLS, the China Health Retirement Longitudinal Study; BMI, body mass index.*

***Table S17: Individual-characteristics with cognitive function in the LASI widowed participants***

|  | **Events/Observations** | **Orientation** | **Memory** | **Executive function** | **Language** |
| --- | --- | --- | --- | --- | --- |
| **Educational attainment** | | | | | |
| Less than upper secondary | 1129/1313 | 0 [Reference] | 0 [Reference] | 0 [Reference] | 0 [Reference] |
| Upper secondary or vocational training | 165/1313 | 0.68(0.53, 0.83) | 0.81(0.67, 0.96) | 0.98(0.84, 1.12) | 0.53(0.36, 0.69) |
| Tertiary | 19/1313 | 0.75(0.35, 1.14) | 0.71(0.32, 1.09) | 1.64(1.27, 2.01) | 0.48(0.05, 0.91) |
| **Household wealth level** | | | | | |
| Low | 475/1313 | 0 [Reference] | 0 [Reference] | 0 [Reference] | 0 [Reference] |
| Normal | 449/1313 | 0.12(0.02, 0.23) | -0.02(-0.13, 0.08) | -0.01(-0.12, 0.09) | 0.04(-0.08, 0.16) |
| High | 389/1313 | 0.06(-0.05, 0.18) | -0.08(-0.19, 0.04) | 0(-0.10, 0.11) | -0.02(-0.15, 0.10) |
| **BMI** | | | | | |
| Underweight/Normal | 928/1313 | 0 [Reference] | 0 [Reference] | 0 [Reference] | 0 [Reference] |
| Overweight | 291/1313 | 0.29(0.17, 0.40) | 0.24(0.12, 0.35) | 0.15(0.04, 0.26) | 0.17(0.04, 0.30) |
| Obesity | 94/1313 | 0.26(0.07, 0.45) | 0.35(0.17, 0.54) | 0.27(0.09, 0.44) | 0.23(0.02, 0.44) |
| **Smoking status** | | | | | |
| Current | 373/1313 | 0 [Reference] | 0 [Reference] | 0 [Reference] | 0 [Reference] |
| Former | 52/1313 | -0.02(-0.27, 0.22) | 0.15(-0.10, 0.39) | 0.01(-0.23, 0.24) | 0.09(-0.18, 0.36) |
| Never | 888/1313 | 0.10(-0.01, 0.21) | 0.06(-0.05, 0.17) | 0.14(0.04, 0.24) | 0.07(-0.05, 0.19) |
| **Drinking patterns** | | | | | |
| Heavy | 11/1313 | 0 [Reference] | 0 [Reference] | 0 [Reference] | 0 [Reference] |
| Moderate | 10/1313 | -0.03(-0.76, 0.69) | -0.16(-0.87, 0.55) | 0.34(-0.34, 1.01) | -0.07(-0.86, 0.72) |
| Light | 83/1313 | -0.22(-0.78, 0.33) | -0.14(-0.69, 0.40) | 0.28(-0.24, 0.80) | -0.31(-0.92, 0.29) |
| Never | 1209/1313 | -0.20(-0.73, 0.33) | -0.02(-0.54, 0.50) | 0.23(-0.26, 0.73) | -0.20(-0.78, 0.38) |
| **Internet use** | | | | | |
| No | 1297/1313 | 0 [Reference] | 0 [Reference] | 0 [Reference] | 0 [Reference] |
| Yes | 16/1313 | 0.11(-0.31, 0.53) | 0.08(-0.34, 0.49) | 0.23(-0.16, 0.63) | 0.27(-0.19, 0.73) |
| **Comorbidities** | | | | | |
| Mild | 1123/1313 | 0 [Reference] | 0 [Reference] | 0 [Reference] | 0 [Reference] |
| Moderate | 171/1313 | 0.11(-0.03, 0.25) | 0.16(0.03, 0.30) | 0.03(-0.10, 0.16) | 0.12(-0.03, 0.27) |
| Severe | 19/1313 | 0.19(-0.20, 0.57) | 0.50(0.12, 0.88) | 0.40(0.04, 0.76) | 0.32(-0.10, 0.74) |

*LASI, the Longitudinal Aging Study in India; BMI, body mass index.*

***Table S18: Individual-characteristics with cognitive function in the HAALSI widowed participants***

|  | **Events/Observations** | **Orientation** | **Memory** | **Executive function** | **Language** |
| --- | --- | --- | --- | --- | --- |
| **Educational attainment** | | | | | |
| Less than upper secondary | 289/307 | 0 [Reference] | 0 [Reference] | 0 [Reference] | 0 [Reference] |
| Upper secondary or vocational training | 16/307 | 0.49(0.02, 0.96) | 0.19(-0.25, 0.64) | 0.34(-0.12, 0.80) | 0.36(-0.11, 0.83) |
| Tertiary | 2/307 | 0.69(-0.62, 1.99) | 1.23(0, 2.47) | 1.33(0.06, 2.61) | 0.39(-0.92, 1.70) |
| **Household wealth level** | | | | | |
| Low | 119/307 | 0 [Reference] | 0 [Reference] | 0 [Reference] | 0 [Reference] |
| Normal | 99/307 | 0.14(-0.11, 0.39) | -0.06(-0.29, 0.18) | 0.10(-0.15, 0.34) | -0.06(-0.31, 0.19) |
| High | 89/307 | 0.15(-0.11, 0.41) | -0.03(-0.27, 0.22) | 0.05(-0.21, 0.30) | -0.27(-0.54, -0.01) |
| **BMI** | | | | | |
| Underweight/Normal | 123/307 | 0 [Reference] | 0 [Reference] | 0 [Reference] | 0 [Reference] |
| Overweight | 95/307 | 0.03(-0.23, 0.29) | -0.18(-0.42, 0.07) | -0.01(-0.26, 0.24) | 0.20(-0.06, 0.46) |
| Obesity | 89/307 | 0.10(-0.17, 0.37) | 0(-0.26, 0.26) | 0.24(-0.03, 0.50) | 0.27(0, 0.55) |
| **Smoking status** | | | | | |
| Current | 6/307 | 0 [Reference] | 0 [Reference] | 0 [Reference] | 0 [Reference] |
| Former | 13/307 | -0.80(-1.80, 0.20) | -1.47(-2.41, -0.52) | -0.79(-1.77, 0.19) | -0.94(-1.94, 0.06) |
| Never | 288/307 | -0.61(-1.53, 0.30) | -0.84(-1.70, 0.02) | -1.06(-1.95, -0.17) | -0.84(-1.75, 0.07) |
| **Drinking patterns** | | | | | |
| Heavy | 11/307 | 0 [Reference] | 0 [Reference] | 0 [Reference] | 0 [Reference] |
| Moderate | 10/307 | 0.10(-0.77, 0.96) | 0.48(-0.34, 1.29) | 0.26(-0.58, 1.10) | 0.83(-0.04, 1.69) |
| Light | 100/307 | -0.21(-0.86, 0.45) | 0.31(-0.31, 0.93) | 0.03(-0.61, 0.67) | 0.42(-0.24, 1.08) |
| Never | 186/307 | 0.10(-0.55, 0.75) | 0.44(-0.17, 1.05) | 0.12(-0.52, 0.75) | 0.54(-0.11, 1.19) |
| **Internet use** | | | | | |
| No | 50/307 | 0 [Reference] | 0 [Reference] | 0 [Reference] | 0 [Reference] |
| Yes | 257/307 | 0.25(-0.04, 0.54) | 0.41(0.14, 0.68) | 0.18(-0.10, 0.46) | 0.62(0.33, 0.91) |
| **Comorbidities** | | | | | |
| Mild | 258/307 | 0 [Reference] | 0 [Reference] | 0 [Reference] | 0 [Reference] |
| Moderate | 43/307 | 0.14(-0.17, 0.44) | 0.12(-0.17, 0.40) | 0.40(0.11, 0.70) | 0.01(-0.30, 0.31) |
| Severe | 6/307 | -0.21(-0.96, 0.55) | -0.05(-0.76, 0.66) | -0.32(-1.06, 0.42) | -0.20(-0.95, 0.56) |

*HAALSI, the Health and Aging in Africa: A Longitudinal Study of an INDEPTH Community in South Africa; BMI, body mass index.*

**Ⅸ. Sensitivity analyses: complete-case analysis**

***Table S19: Association of between spousal loss and cognitive function using complete-case design***

|  | **HRS (n=2034)** | **ELSA (n=911)** | **MHAS (n=1116)** | **CHARLS (n=4000)** | **LASI (n= 3349)** | **HAALSI (n=295)** |
| --- | --- | --- | --- | --- | --- | --- |
| **Orientation** | 0.03(-0.07, 0.14) | 0.15(-0.02, 0.32) | -0.06(-0.20, 0.09) | -0.12(-0.21, -0.03) | -0.09(-0.15, -0.02) | -0.13(-0.37, 0.11) |
| **Memory** | -0.03(-0.12, 0.06) | 0.08(-0.05, 0.22) | -0.17(-0.29, -0.04) | -0.15(-0.24, -0.06) | -0.04(-0.10, 0.03) | -0.16(-0.40, 0.07) |
| **Executive function** | -0.04(-0.12, 0.04) | 0.01(-0.12, 0.15) | -0.06(-0.17, 0.05) | -0.20(-0.29, -0.10) | -0.020(-0.08, 0.04) | -0.13(-0.37, 0.12) |
| **Language** | -0.02(-0.11, 0.08) | 0.13(-0.02, 0.29) | 0.02(-0.11, 0.15) | -0.16(-0.25, -0.07) | -0.04(-0.11, 0.04) | -0.19(-0.45, 0.08) |

*HRS, the Health and Retirement Study; ELSA, the English Longitudinal Study of Ageing; MHAS, the Mexican Health and Aging Study; CHARLS, the China Health Retirement Longitudinal Study; LASI, the Longitudinal Aging Study in India; HAALSI, the Health and Aging in Africa: A Longitudinal Study of an INDEPTH Community in South Africa.*

**Ⅹ. Sensitivity analyses: baseline-exposure analysis**

***Table S20: Association of between spousal loss and cognitive function using a design that includes the participants only from baseline wave***

|  | **HRS (n=2528)** | **ELSA (n=1033)** | **MHAS (n=1451)** | **CHARLS (n=6981)** | **LASI ^c^ (n=3924)** | **HAALSI (n=540)** |
| --- | --- | --- | --- | --- | --- | --- |
| **Orientation** | -0.02(-0.04, 0.00) | 0.04(0.01, 0.08) | -0.07(-0.10, -0.04) | -0.07(-0.09, -0.06) | -0.02(-0.04, 0.01) | 0.02(-0.02, 0.06) |
| **Memory** | -0.08(-0.09, -0.06) | -0.02(-0.05, 0.01) | -0.14(-0.17, -0.12) | -0.12(-0.14, -0.11) | -0.04(-0.07, -0.02) | -0.08(-0.12, -0.04) |
| **Executive function** | -0.10(-0.11, -0.08) | -0.02(-0.05, 0.01) | -0.08(-0.10, -0.06) | -0.13(-0.15, -0.12) | -0.05(-0.08, -0.03) | -0.15(-0.19, -0.11) |
| **Language** | -0.04(-0.06, -0.02) | 0.07(0.03, 0.10) | -0.02(-0.04, 0.01) | -0.17(-0.19, -0.15) | -0.05(-0.08, -0.02) | -0.14(-0.18, -0.10) |

*Note: c means that LASI just releases the first wave data and the LASI β (95% CI) shown is from the main analysis.*

*HRS, the Health and Retirement Study; ELSA, the English Longitudinal Study of Ageing; MHAS, the Mexican Health and Aging Study; CHARLS, the China Health Retirement Longitudinal Study; LASI, the Longitudinal Aging Study in India; HAALSI, the Health and Aging in Africa: A Longitudinal Study of an INDEPTH Community in South Africa.*

**Ⅺ. Sensitivity analyses: age-restriction analysis**

***Table S21: Association of between spousal loss and cognitive function using a design that restricts the HCAP participants age ≥ 65 years***

|  | **HRS ^d^ (n=2672)** | **ELSA ^d^ (n=1075)** | **MHAS (n=1009)** | **CHARLS (n=5879)** | **LASI (n=2826)** | **HAALSI (n=417)** |
| --- | --- | --- | --- | --- | --- | --- |
| **Orientation** | 0.00(-0.02, 0.02) | 0.10(0.06, 0.13) | -0.09(-0.12, -0.06) | -0.08(-0.09, -0.06) | -0.05(-0.07, -0.04) | 0.08(0.03, 0.13) |
| **Memory** | -0.06(-0.07, -0.04) | 0.05(0.02, 0.08) | -0.11(-0.14, -0.08) | -0.10(-0.11, -0.08) | -0.03(-0.05, -0.02) | -0.14(-0.19, -0.09) |
| **Executive function** | -0.09(-0.10, -0.07) | 0.01(-0.02, 0.03) | -0.07(-0.10, -0.05) | -0.13(-0.14, -0.12) | 0.01(0.00, 0.03) | -0.17(-0.22, -0.12) |
| **Language** | -0.05(-0.07, -0.04) | 0.16(0.12, 0.19) | 0.04(0.01, 0.07) | -0.16(-0.17, -0.14) | 0.01(-0.01, 0.03) | -0.05(-0.10, 0.00) |

*Note: d means the HRS-HCAP and ELSA-HCAP includes participants aged 65 years and older, and the HRS and ELSA β (95% CI) shown is from the main analysis.*

*HRS, the Health and Retirement Study; ELSA, the English Longitudinal Study of Ageing; MHAS, the Mexican Health and Aging Study; CHARLS, the China Health Retirement Longitudinal Study; LASI, the Longitudinal Aging Study in India; HAALSI, the Health and Aging in Africa: A Longitudinal Study of an INDEPTH Community in South Africa.*

**Ⅻ. Sensitivity analyses: excluding high-income national cohorts analysis**

***Table S22: Associations between spousal loss and subsequent domain-specific cognitive function (Excluding HRS and ELSA)***

|  | ***β* (95% CI)** | ***p*-value** | ***I*^2^** |
| --- | --- | --- | --- |
| **Orientation** | -0.08 (-0.09, -0.07) | <0.001 | 1.97% |
| **Memory** | -0.09(-0.14, -0.05) | <0.001 | 96.67% |
| **Executive function** | -0.10(-0.17, -0.03) | 0.0034 | 98.38% |
| **Language** | -0.07(-0.14, 0.00) | 0.0375 | 98.41% |

*95% CI: 95% confidence interval.*

*HRS, the Health and Retirement Study; ELSA, the English Longitudinal Study of Ageing.*
